# Urine extracellular vesicles capture kidney transcriptome and hyperglycemia linked mRNA signatures for type 1 diabetic kidney disease

**DOI:** 10.1101/2021.10.11.21264829

**Authors:** Om Prakash Dwivedi, Karina Barreiro, Annemari Käräjämäki, Erkka Valo, Rashmi B Prasad, Harry Holthöfer, Per-Henrik Groop, Tiinamaija Tuomi, Carol Forsblom, Leif Groop, Maija Puhka

## Abstract

Diabetic kidney disease (DKD) is a severe complication of type 1 diabetes (T1D), which lacks non-invasive early biomarkers. Although less explored, mRNAs in urinary extracellular vesicles (uEV) could reflect changes in the kidney transcriptome during DKD development. We performed genome-wide mRNA sequencing of >100 uEV samples from two T1D cohorts with 24-hour and overnight urine collections. Our uEV pipeline allowed reproducible detection of >10,000 mRNAs bearing overall similarity to kidney transcriptome. uEV from T1D DKD groups showed significant upregulation of 13 genes, prevalently expressed by proximal tubular cells within the kidney. Strikingly, six genes involved in cellular stress responses including protection against oxidative stress (*GPX3, NOX4, MSRB, MSRA, HRSP12* and *CRYAB*) correlated with hyperglycemia and long-term changes in kidney function independent of albuminuria status. The study identified genes associated with glycemic stress in T1D DKD and confirmed the utility of uEV in capturing pathological gene expression signatures from kidney.

## Introduction

Diabetes is a multifactorial disease with diverse complications causing a global health burden and >1.3 million annual deaths (*1*). One of the most severe complications is diabetic kidney disease (DKD) - a leading cause of end-stage renal disease (*2*). At tissue level, DKD is characterized by histological features such as glomerular basement membrane thickening, mesangial matrix expansion, nodular glomerulosclerosis, and arteriolar hyalinosis – more often detected in type 1 diabetes (T1D) compared to type 2 diabetes (T2D) (*2-4*). Kidney biopsy is considered as the gold standard to observe the DKD linked structural changes and progression stages (*5*). However, in practice, the diagnosis is usually based on albuminuria and decline in kidney function measured as glomerular filtration rate (*6, 7*). Despite their wide clinical applications, these traditional markers are non-specific and poorly detect early stages of DKD, for example hyperfiltration (*8*).

Transcriptomics of the kidney tissue has suggested association of kidney mRNA markers with DKD related structural changes like fibrosis (*9, 10*), and also led to development of sensitive biomarkers like epidermal growth factor (*11*). However, the invasiveness and rarity of biopsies has limited kidney transcriptome-based approaches in DKD precision medicine (*12*). This has turned the interest to extracellular RNAs in urine, which might mirror or provide novel insight into the pathological changes of kidney transcriptome and an alternative non-invasive way for diagnosing, prognostication and monitoring DKD (*13*).

Extracellular vesicles (EV) are lipid bilayer contained particles secreted by cells in varying sizes (∼30-1000 nm). They carry active biomolecules (proteins, lipids, DNA and RNA) and contribute to many biological processes in health and disease e.g. through their role in cell-to-cell signaling (*14-16*). EV are found in all body fluids, where their membrane helps to protect the cargo from proteases and RNases (*17*). Thus, EV provide a potentially stable biomarker reservoir that reflects the status of the cell of origin (*18*).

Due to the rich small RNA content of urinary EV (uEV), DKD research has focused mainly on uEV miRNAs (*19-22*). In contrast, although uEV seem to carry a significant part of the cell free mRNA, studies on mRNA are rare, particularly using non-targeted genome-wide next-generation sequencing (NGS) in any significant number of urine samples (*23-28*). Reasons include missing standards for urine collections and EV isolation methods (*29*), and the need for optimized low-input NGS protocols. Recently, we successfully tested such methods for uEV mRNA profiling (*28*), but information of their reproducibility and applicability in larger cohorts of T1D DKD patients is still lacking.

In this study, we utilized low input mRNA sequencing (mRNAseq) for uEV and analyzed the whole uEV mRNA transcriptome in >100 samples including T1D individuals with varying stages of DKD (i.e. normo-, micro- or macroalbuminuria). We show 1) technical reproducibility of the uEV mRNA sequencing pipeline and feasibility in clinical setting by combining cohorts with different urine collection protocols, 2) comprehensive characterization of uEV mRNA transcriptomes with respect to other human tissues suggesting its similarity with kidney, and 3) identification of novel candidate marker genes that may have potential role in maintaining cellular stress response in kidney during DKD pathogenesis in T1D individuals.

## Results

### Study design

The uEV mRNAseq study of 102 samples focused on three main aims: 1) to assess the feasibility and reproducibility of low input uEV mRNAseq for different types of clinical urine collections, 2) to assess how uEV capture gene expression signatures of the kidney or other tissues, and 3) to discover novel candidate genes for kidney disease in individuals with T1D (**Fig. 1**). The tested technical variables were: 1) overnight (ON) vs. 24-hour (24-h) urine collection (n=6 donors, 12 pairs), 2) urine samples processed with or without centrifugation prior to freezing (n=4 donors, 4 pairs), and 3) technical replicates of urine passing through the whole pipeline from uEV isolation to mRNAseq at 1-5 months intervals (n=8 donors, 6 duplicates and 2 triplicates) (**Fig. 1-**technical comparisons). To find novel DKD candidate genes, we performed mRNAseq for uEV samples from 72 T1D individuals collected from two different T1D cohorts (FinnDiane, Finnish Diabetic Nephropathy Study; n=34 and DIREVA, Diabetes Registry in Vaasa; n=38) with different urine collection types (24-h and ON urine collection, respectively) (**Fig. 1**). The included samples differed regarding the degree of urine albuminuria (normoalbuminuria, n=38; microalbuminuria, n=15 and macroalbuminuria, n=19) as well as estimated glomerular filtration rate (eGFR), HbA_1c_ (glycated hemoglobin A1c) and diabetes duration (table S1, S2, S3). We also utilized the long-term (15 years) clinical laboratory data (eGFR, HbA_1c_) for association analysis. As the study included only men to reduce heterogeneity (e.g. due to differences in the genitourinary system), we assessed the impact of sex on the expression of all robustly detected uEV mRNAs by clustering analysis (women, n=3, men, n=3 or 67) (**Fig. 1**).

**Fig. 1:**
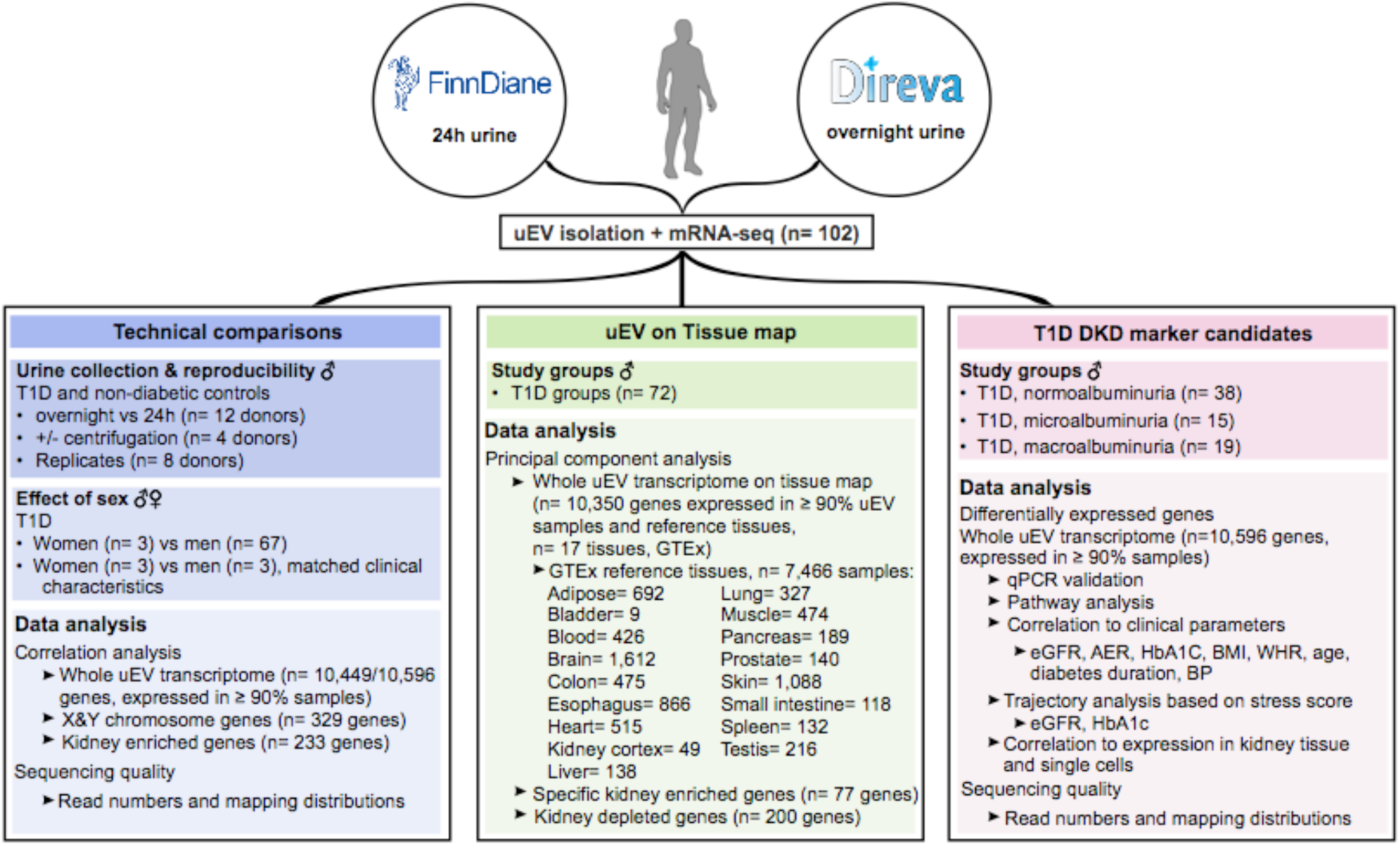
Study design. Figure depicting study design and work flow for next generation sequencing of uEV mRNAs and discovery of candidate markers for T1D DKD. 24 hour (24h); albumin excretion rate (AER); blood pressure (BP); body mass index (BMI); diabetic kidney disease (DKD); estimated glomerular filtration rate (eGFR); Genotype-Tissue Expression (GTEx) project (30); glycated hemoglobin (HbA1_C_); macroalbuminuria (macro); microalbuminuria (micro); normoalbuminuria (normo); messenger RNA sequencing (mRNAseq); principal component analysis (PCA); proximal convoluted tubule (PCT); quantitative polymerase chain reaction (qPCR); type 1 diabetes (T1D); urinary extracellular vesicles (uEV); waist-to-hip ratio (WHR).

Additionally, we combined the mRNA expression profiles of uEV and various tissues obtained from the Genotype-Tissue Expression (GTEx) project (*30, 31*) (17 tissues, n=7,466 tissue samples) to assess the similarity of uEV mRNAs to mRNAs from kidney cortex and other tissues (**Fig. 1**, tissue map).

### Quality control of uEV

Urinary EV isolates obtained by ultracentrifugation from the different study groups produced acceptable uEV and RNA quality, as shown by electron microscopy (EM), Western blotting, nanoparticle tracking analysis (NTA) and EV-RNA profiling by Bioanalyzer Pico assay (**Fig. 2**). We detected typical round EVs showing different staining intensities, sizes and morphologies by EM (**Fig. 2, A** and **B**). The isolates contained also some Tamm-Horsfall protein filaments. The EVs were roughly up to 1 *µ*m in diameter, although the main population consisted of smaller (exosomal range) sizes (30-150 nm). NTA indicated a similar size range of particles (**Fig. 2C)**. Western blotting showed variable quantities of two uEV-enriched proteins, CD9 and podocalyxin, in the study samples (**Fig. 2D)**. Despite the variable EV marker quantities, sufficient amounts of total RNA could be extracted from all EV samples (mean 22 ng, range 2-140 ng). Bioanalyzer showed typical RNA profiles for the uEV from all study groups: a relatively large amount of small RNA and no or minor peaks of rRNA (**Fig. 2E**). We have already previously found the uEV quality from this ultracentrifugation protocol to be good after a thorough characterization of a set of 10 samples comprising 24-h urines from healthy controls and individuals with T1D (*28*)-these samples were included in both this study and in Barreiro et al. (*28*).

**Fig. 2:**
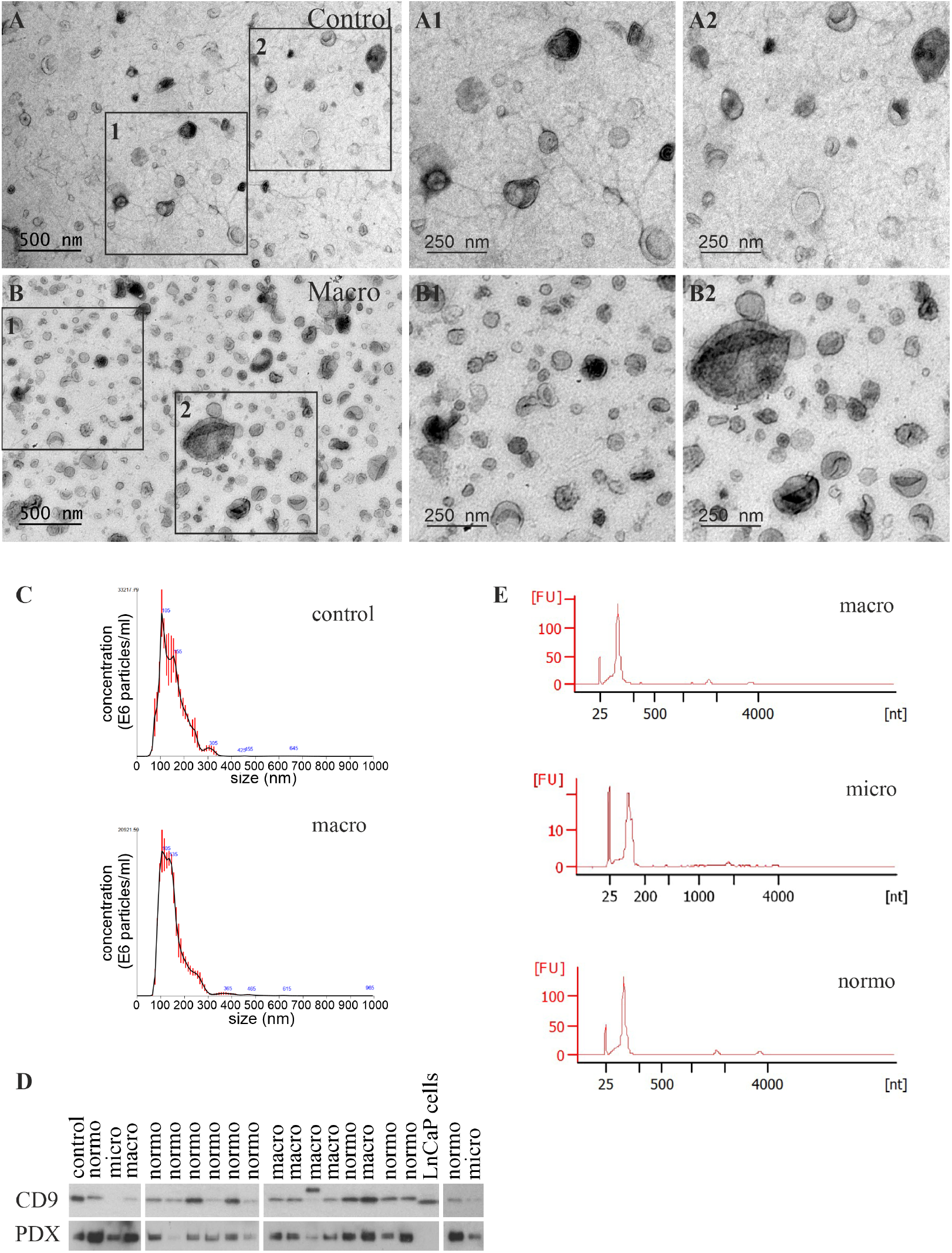
Quality control of the uEV preparations. **(A** and **B)**, Transmission electron micrographs showing uEV of typical morphology and variable sizes; filamentous structures compatible with Tamm–Horsfall protein filaments are present in the in-set **A1. (C)**, Representative NTA profiles showing enrichment of vesicle sizes between 50-300 nm. (**D)**, Western blotting showing the presence of uEV-enriched markers CD9 and PDX in uEV preparations. (**E)**, Representative electropherograms of RNA isolated from uEV and analyzed with Bioanalyzer using Agilent Pico kit. A typical RNA peak between 25-200 nt was detected in all samples. Lymph Node Carcinoma of the prostate (LnCaP), macroalbuminuria (macro); microalbuminuria (micro); nanoparticle tracking analysis (NTA);gene expression data for all commonly expressed non-diabetic control (control); normoalbuminuria (normo); Podocalyxin (PDX), urinary extracellular vesicles (uEV).

### Quality, robustness and reproducibility of uEV mRNA profiling by NGS

We performed NGS of uEV mRNAs using low RNA input protocol for both T1D cohorts (total=72, 24-h or ON urine collection) and three different technical comparisons (**Fig. 1**). In the T1D cohorts, we observed a median (25% - 75% percentile) of 11.5 million (7.4-16.3) raw sequencing reads (fig. S1A). However, the read distributions differed between the cohorts - the ON tended to yield higher read numbers than the 24-h collections. Most raw sequencing reads (median% [25%-75% percentile], 90.4% [87.6-91.9]) mapped uniquely to the human genome with <7% of unmapped or multi-mapped reads (4.6% [3.7-5.9] and 4.7% [3.7-6.5], respectively) (fig. S2). Furthermore, as expected, most uniquely mapped reads aligned to exonic genomic regions (73.6% [72.3-74.6] for CDS exons, 14.5% [13.7-15.7] for 3’UTR exons and 11.6% [10.9-12.8] for 5’UTR exons), and <0.1% mapped to intronic or intergenic regions (10 Kb upstream or downstream of coding regions) (fig. S3).

We detected robust expression of 10,596 protein coding genes in uEV from the T1D cohorts that were expressed in >90% of samples (with count ≥1). The distribution of gene expression of all the genes appeared similar for both T1D cohorts despite the differences in urine collection protocols (**Fig. 3A)**. To further test for differences in uEV mRNA profiles, we performed principal component analysis (PCA) of all (n=10,596) robustly expressed genes. We observed no systematic differences as 67 out of 72 samples clustered together (**Fig. 3B**). However, the five outliers were removed from downstream case-control analysis.

**Fig. 3:**
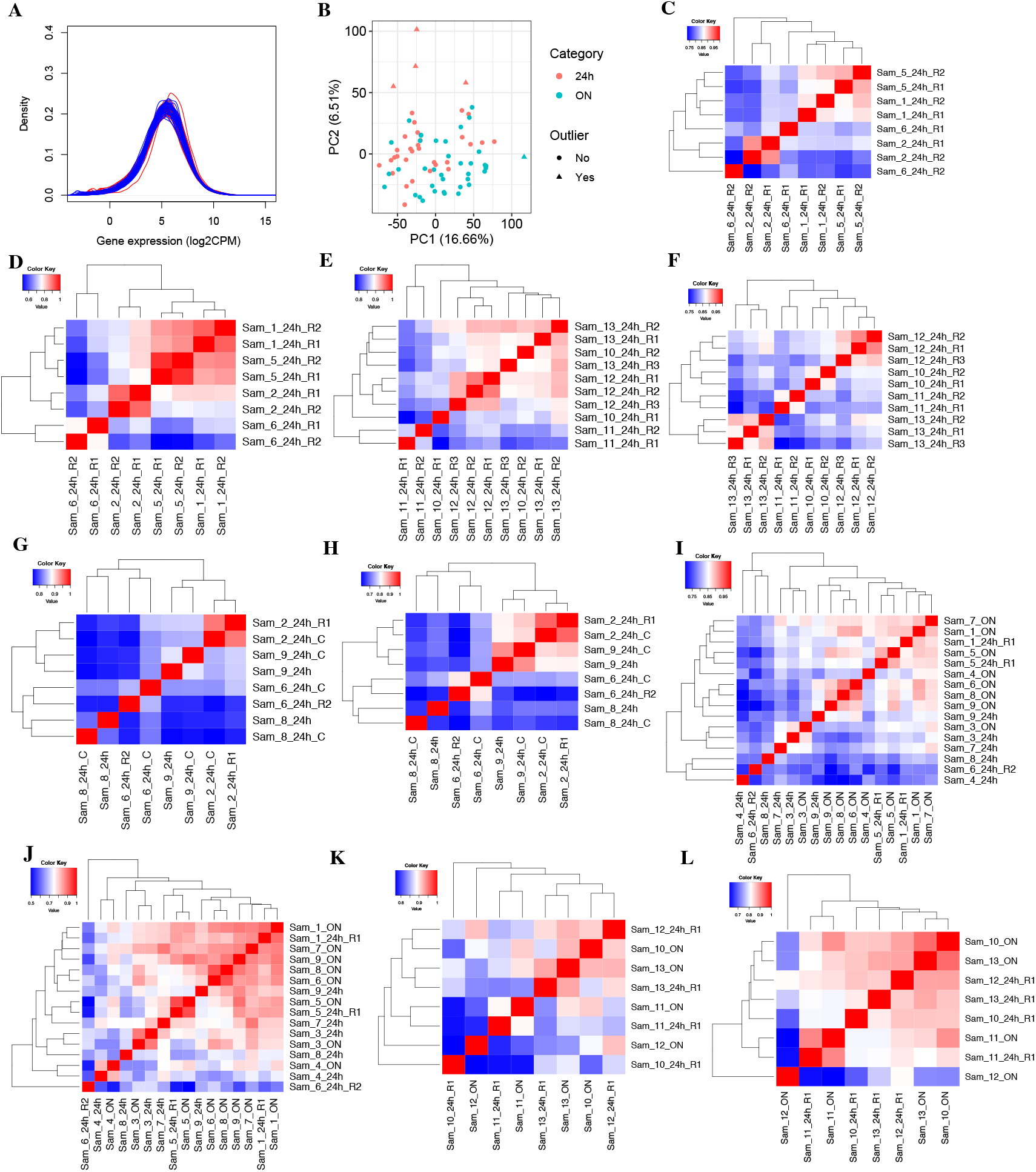
Sequencing of uEV mRNAs showed consistent gene expression distribution and reproducibility across different sample sets. **(A)**, Density plot depicting gene expression distribution (log2CPM) for all commonly expressed protein coding genes (count ≥1 in ≥90% samples, n=10,596) in urine samples from the T1D cohorts (total n=72; 24h (24 hour) collections in blue, n=34; ON (overnight) collections in red, n=38). (**B)**, Principal component (PC) analysis using gene expression data for all commonly expressed genes (n=10,596) in the T1D cohorts (n=72). **(C** to **L)**, Hierarchical clustering based on gene expression correlation matrix (Spearman’s) using all commonly expressed genes (n=10,449, in **C, E, G, I** and **K**) or kidney enriched genes (n=233, in **D, F, H, J** and **L**) for the sample sets included in technical comparisons: (**C** to **F)**, Technical replicates (**C**-**D**, T1D, and **E**-**F**, non-diabetic donors). Aliquots of individual samples (24h urine collection) were processed through the uEV mRNA sequencing pipeline twice (R1 and R2, n=6 donors, T1D: Samples 1, 2, 5 and 6; Non-diabetic: Samples 10 and 11) or thrice (R1, R2 and R3, n=2 non-diabetic donors: Samples 12 and 13) at different time points. (**G** and **H)**, Centrifugation test samples (T1D donors). Urine samples (24h collection) were processed with (_C) and without centrifugation before freezing (n=4 T1D donors: Samples 2, 6, 8 and 9). (**I** to **L)**, Samples from ON vs 24h urine collections (**I**-**J**, T1D, and **K**-**L**, non-diabetic donors). Samples from individual donors were collected from the same day (n=12 donors, T1D: Samples 1, 3-9; Non-diabetic: Samples 10-13). The gene expression values (log2CPM) were inverse normally transformed and converted to z score unit (sd unit) before analysis. Counts per million (CPM); principal component (PC); replicate (R); sample (Sam); type 1 diabetes (T1D); urinary extracellular vesicles (uEV).

We further tested the robustness of uEV mRNA profiles using pairs of samples for three different technical variables, 1) urine collection protocol: 24-h vs. ON, 2) effect of centrifuging the urine samples prior to freezing (± centrifugation), and 3) reproducibility by processing aliquoted urine samples through the whole uEV mRNAseq pipeline - from uEV isolation to sequencing - at different time points (replicates) (**Fig. 1** and table S4). In these three technical comparisons, similar to the main T1D study cohorts, we observed (median-IQR) 11.5 (8.6-13.5) million raw sequencing reads that were mostly [91.6% (88.2-93.9)] uniquely mapped to human genome, particularly to the exonic gene regions compared to intronic or non-coding regions (fig. S1B, S4A and S4B). We detected the expression of 10,449 protein coding genes (with count ≥1) present in ≥ 90% of samples used for the three technical comparisons. The read count data for these commonly expressed genes (10,449) showed clustering (based on correlation) for most technical replicates (fig. S4C). We performed hierarchical clustering of the sample pairs based on correlation of gene expression using all common detected protein coding genes (n=10,449) or “kidney enriched genes” (n=233, present in uEV out of 413) – a subset of protein coding genes known to be enriched in kidney (see Methods) and expressed in our technical uEV sample sets. Pairs of replicate urine samples and those with or without centrifugation showed high correlation (r=0.81 to 0.95) (fig. S5) and most of these technical pairs clustered together both when all (**Fig. 3, C, E** and **G**) or when kidney enriched genes were considered **(Fig. 3, D, F** and **H**). All 24-h and ON pairs did not cluster together but no systematic difference was observed for the urine sample pairs either from T1D individuals or non-diabetic individuals regarding all commonly expressed genes (**Fig. 3, I** and **K**) or kidney enriched genes (**Fig. 3, J** and **l**). Finally, we also tested sex-dependent differences in uEV mRNA profiles by hierarchical clustering of three samples from women with either three samples from men matched for the stage of albuminuria (n=3) or including all the men from the T1D cohorts (n=67). Men and women clustered separately based on global gene expression (n=10,596 genes) and sex chromosome genes (n=329), but not based on kidney enriched genes (n=247 present in uEV out of 413, see methods) (fig. S6 and S7).

Together, these results showed the expression of >10,000 protein coding genes in uEV from different clinical cohorts. It is noteworthy that this is the first study that successfully combined a relatively large number of samples from different urine collections highlighting the comparability and reproducibility of the uEV mRNA profile and the used technologies.

### Similarity of the uEV mRNA profile with kidney transcriptome

Gene expression profiles of uEV could be derived from several tissues – thus, to study the origin of the uEV mRNAs, we compared the uEV transcriptome to individual organ -specific gene expression signatures. We obtained sets of known “kidney enriched genes” (413 genes with higher expression in kidney than in other tissues, see the Methods) and “kidney depleted genes” (4,631 genes not detected in kidney) from the human protein atlas database (*32, 33*), and checked their expression in T1D uEV (with the total of n=10,596 genes). We detected the expression of 59.8% (247 out of 413) of kidney enriched genes compared to only 4.3% (201 out of 4,631) of kidney depleted genes in the T1D uEV. We further looked for more “specific kidney enriched genes”-a subset of kidney enriched genes that are reported to be detected in kidney and only in some human tissues (see Methods)- and detected the expression of 40.5% of such genes (77 out of 190) (**Fig. 4A**).

**Fig. 4:**
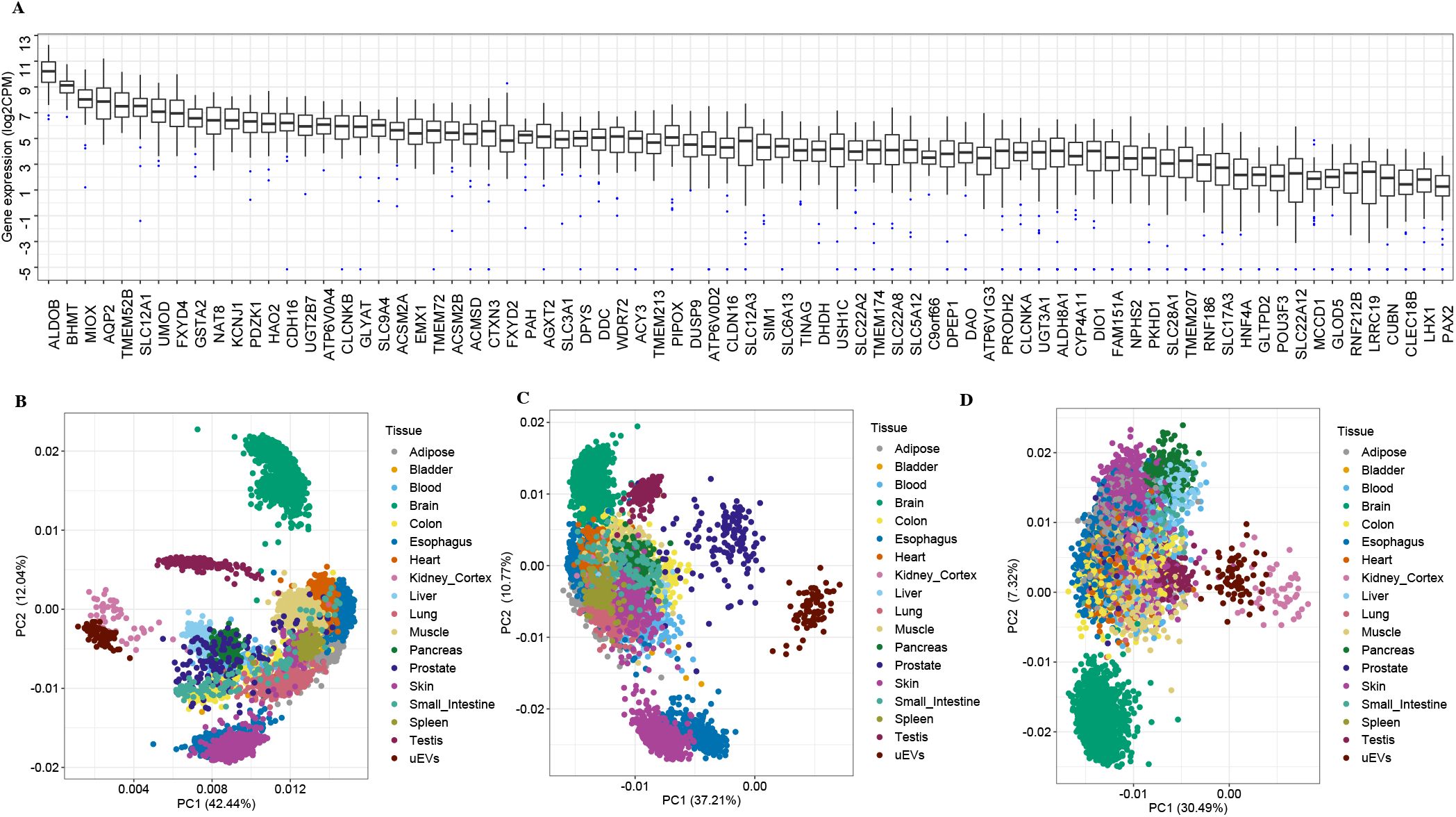
Genome-wide mRNA profile of uEV showed similarity with the kidney mRNA profile. **(A)**, The boxplot depicts the interquartile range, median and minimum/maximum summary values of expression levels of the 77 known specific kidney enriched genes that are reported to be detected in kidney and only in some human tissues (see Methods). (**B** to **D)**, PC analysis using gene expression levels (log2CPM) in uEV from the T1D cohorts (n=72), and the respective gene expression levels (log2CPM) in 17 human tissues obtained from the GTEx reference database (*30, 31*) (tissues from men only; Adipose=692, Bladder=9, Blood=426, Brain=1612, Colon=475, Esophagus=866, Heart=515, Kidney Cortex=49, Liver=138, Lung=327, Muscle=474, Pancreas=189, Prostate=140, Skin=1088, Small Intestine=118, Spleen=132, Testis=216). (**B)**, all commonly expressed protein coding genes as detected in uEV and GTEx tissues (n=10,350); (**C)**, a subset of “kidney depleted genes”, i.e. genes normally not expressed in kidney (n=200 present in uEV+GTEx combined dataset out of 4,631, details in Methods); (**D), “**specific kidney enriched genes”, a subset of kidney enriched genes (n=77, as shown in **A**) that are reported to be detected in kidney and only in some other human tissues (see Methods). Counts per million (CPM); genotype-Tissue Expression (GTEx); principal component (PC); type 1 diabetes (T1D); urinary extracellular vesicles (uEV).

We next compared the expression profile of uEV mRNAs with the profiles of 17 other human tissues obtained from the GTEx database (*30, 31*) (all male, **Fig. 4**). To infer proximity between the profiles, we performed PCA analysis using expression levels of three sets of genes; 1) all protein coding genes expressed in both uEV and the 17 reference human tissues (n=10,350), 2) a subset of “kidney depleted genes” as negative control and 3) a subset of “specific kidney enriched genes” (n=77, as shown in **Fig. 4A**). PCA analysis using gene expression data of all protein coding genes showed that uEV samples clustered together with the kidney cortex tissue samples and away from other tissues suggesting close similarity of the mRNA expression patterns between uEV and kidney (**Fig. 4B**). The PCA analysis of kidney depleted genes showed proximity of prostate samples with uEV samples (**Fig. 4C**). We observed a similar pattern, clustering of kidney cortex and uEV samples, when using specific kidney enriched genes in the PCA analysis (**Fig. 4D**).

In summary, we detected expression of a high number of kidney enriched genes in uEV and generally a lack of expression of genes not known to be expressed in kidney. The overall expression pattern of protein coding genes in uEV also showed close similarity with kidney suggesting that kidney is the main contributor of mRNAs detected in uEV.

### Genome-wide uEV mRNA analysis revealed novel candidate markers for T1D DKD

We performed hypothesis-free global differential gene expression analysis using mRNA expression profiles of uEV (n=10,596 genes) and comparing macroalbuminuric and normoalbuminuric T1D individuals (n=17 and n=37, respectively, mRNAseq PCA quality passed) (table S2 and **Fig. 5)**. The analysis revealed differential expression of 13 genes (*MAP7, MSRB1, GPX3, IL32, NOX4, HRSP12, TINAG, CAPN3, CXCL14, MSRA, CRYAB, RBP5* and *TMEM9*) with robust significance (P≤ 3.85 10^−6^) after accounting for multiple correction of all genes (**Fig. 5A**). Interestingly, the expression level of all the differentially expressed genes was upregulated in the uEV of macroalbuminuric T1D individuals (**Fig. 5, A** and **B**).

**Fig. 5:**
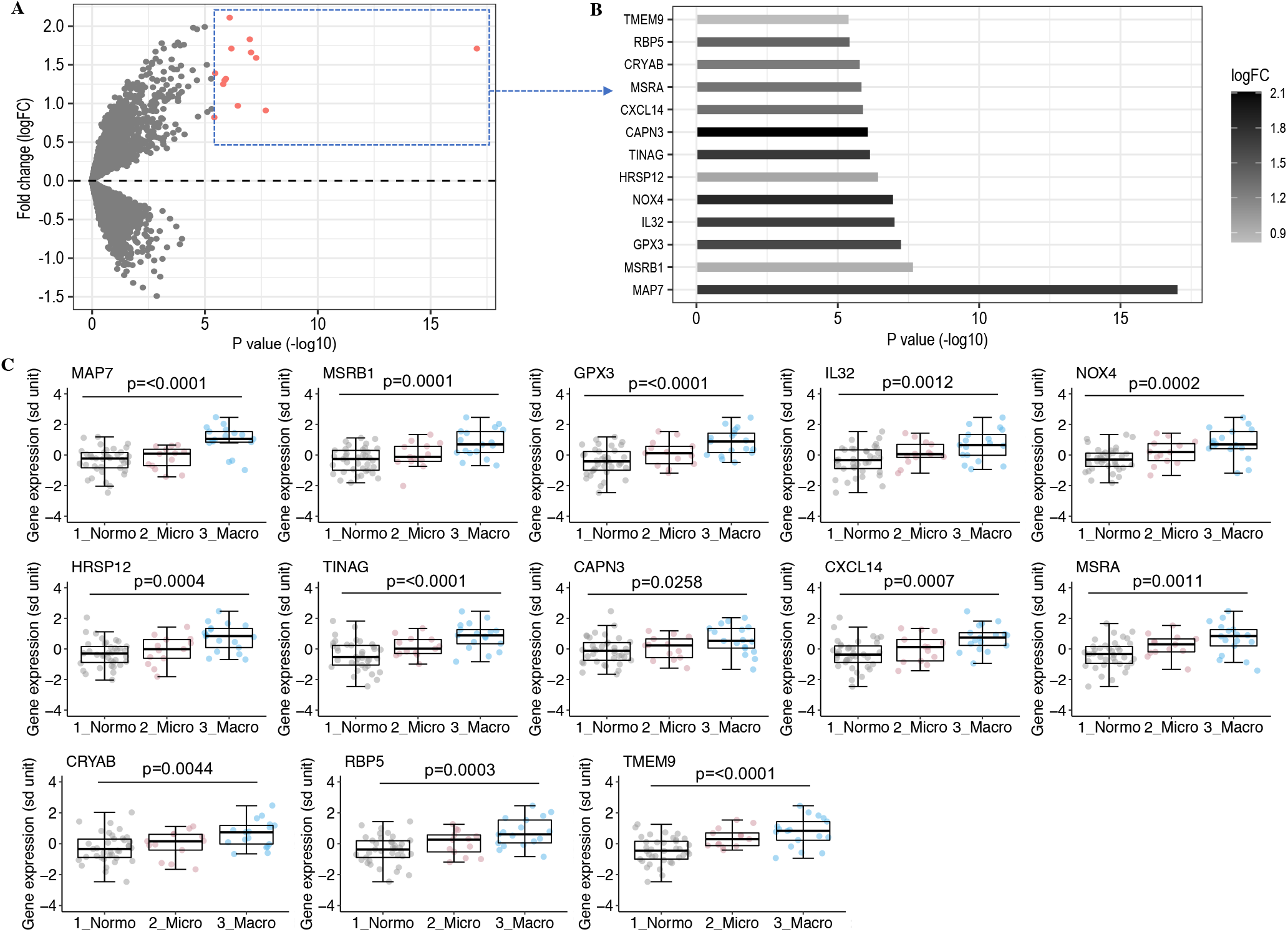
Global differential gene expression analysis revealed upregulation of 13 transcripts in the uEV of type 1 diabetic individuals with macroalbuminuria. **(A** and **B)**, Global DGE analysis (n=10,596 genes) comparing macro- (n=17) and normoalbuminuric T1D (n=37) individuals, depicts significantly (p≤3.85×10^−6^) differentially expressed transcripts (red points in **A**; names and respective logFC in **B**). (**C)**, The box plots illustrate the gene expression distribution (sd unit) of 13 differentially expressed genes in macro- and normoalbuminuric T1D subjects (as in **A**) along with the group of microalbuminuric T1D patients (n=13). The DGE analysis (Normo vs Macro) in **A** and **B** was performed based on count data using generalized linear models adjusting for age, body-mass index, diabetes duration and urine collection protocols (overnight and 24 hour) and multiple correction using the Bonferroni method. In **C**, the three groups comparisons were performed using one-way ANOVA. The gene expression values (log2CPM) were inverse normally transformed and converted to z score unit (sd unit). The boxplots depict the interquartile range, median and minimum/maximum summary values. Differential gene expression (DGE); fold change (FC); log2 normalized counts per million (log2CPM); macroalbuminuria (Macro); microalbuminuria (Micro); normoalbuminuria (Normo); type 1 diabetes (T1D); urinary extracellular veiscles (uEV).

Having shown the difference between normo- and macroalbuminuric individuals, we then included also the microalbuminuric group (n=13, mRNAseq PCA quality passed) to study expression of the identified 13 genes. The gene expression tended to increase from normo- to micro- to macroalbuminuria (**Fig. 5C)**. While the difference between normo- and microalbuminuric individuals was not statistically significant for all the genes, the combined micro- and macroalbuminuria group differed significantly from the normoalbuminuric group (table S5).

Finally, we reanalyzed the uEV mRNA profile of T1D individuals based on CKD stage, irrespective of albuminuria, using retrospective eGFR data (> 5 years) from routine laboratory follow-up testing for CKD classification (table S3). This confirmed significant (p<0.004) upregulation of all 13 differentially expressed genes also in T1D individuals with CKD stage ≥3 compared with CKD stage ≤2 (table S5).

To technically validate the differential expression results from NGS, we studied the expression of five of the top differentially expressed genes (*MAP7, GPX3, IL32, NOX4, HRSP12*) using Taqman qPCR assays in the same set of samples. The qPCR- and mRNAseq-based expression values were mostly highly correlated (**Fig. 6A)**. Similar to the mRNAseq results, the qPCR-based expression levels were significantly (p≤0.005) upregulated in macroalbuminuric T1D individuals and less so in microalbuminuric subjects compared to those with normoalbuminuria (**Fig. 6B**).

**Fig. 6.**
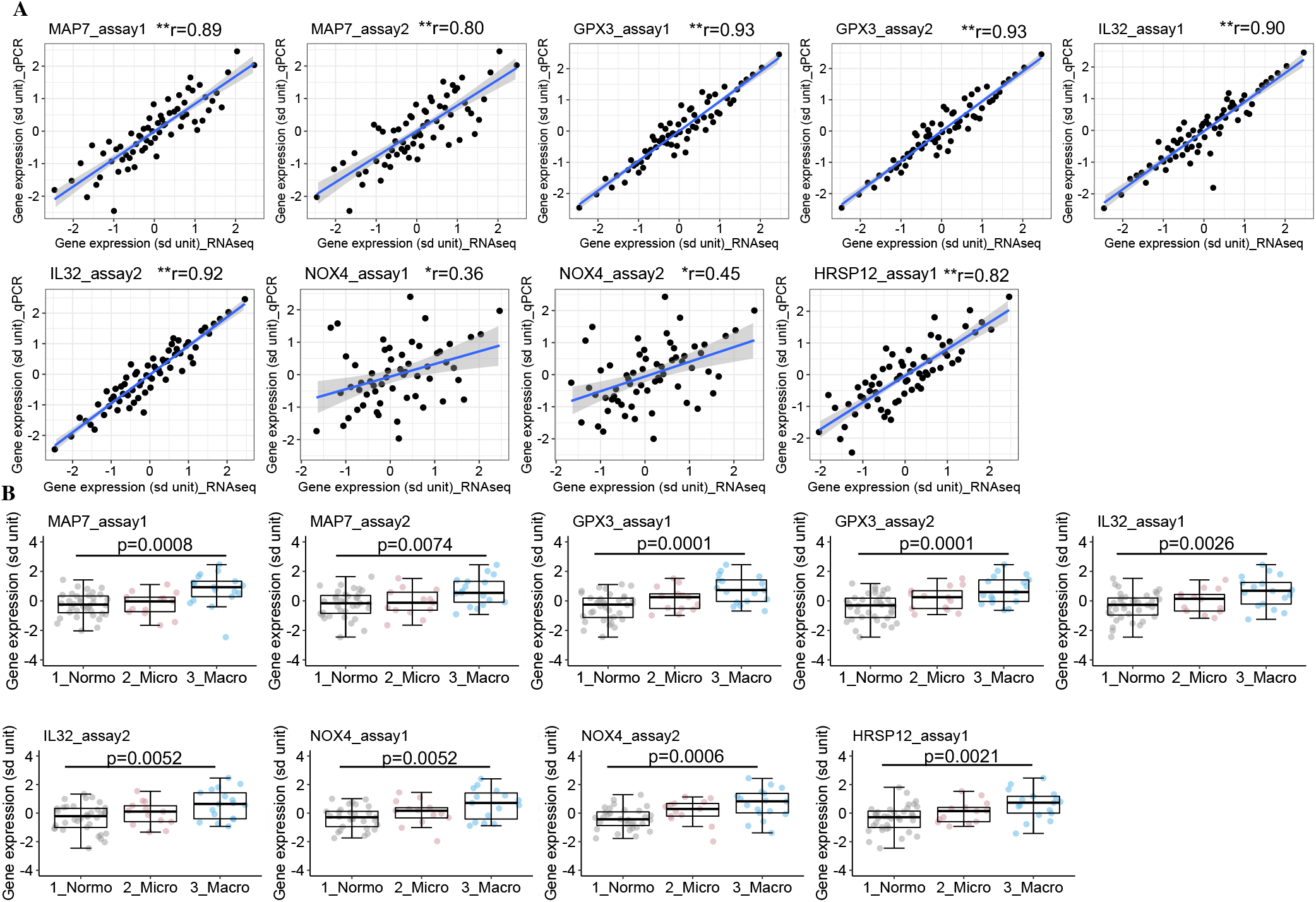
Validation of mRNA sequencing -based differentially expressed genes using quantitative PCR. Differential expression of five of the top 13 DKD associated transcripts of uEV (*MAP7, GPX3, IL32, NOX4, HRSP12*; Fig. 5) was validated by qPCR. (**A)**, Correlation of the gene expression level by next generation mRNA sequencing (x-axis) vs. qPCR (y-axis) in the same samples (n=57-66). DeltaCt values from the qPCR experiments and log2 counts per million values from RNA sequencing experiments were inverse normally transformed and converted to z score unit (sd unit); r=non-parametric Spearman’s (two-tailed) method; *p<0.006, **p<0.00001. (**B)**, The boxplots (interquartile range, median and minimum/maximum summary values) show the gene expression level derived from qPCR assays using 1-2 Taqman assays (assay1 and assay 2) per gene in T1D subjects stratified for the degree of albuminuria (normo- (n=29-36), micro- (n=13), macroalbuminuria (n=15-17). The gene expression values (deltaCt values from qPCR) were inverse normally transformed and converted to z score unit (sd unit) and statistical comparisons of the three groups were performed using one-way ANOVA. Diabetic kidney disease (DKD); macroalbuminuria (Macro); microalbuminuria (Micro); normoalbuminuria (Normo); quantitative PCR (qPCR); type 1 diabetes (T1D); uEV; urinary extracellular vesicles (uEV).

### T1D DKD differential uEV mRNAs correlation with hyperglycemia and co-expression similarity with kidney

We studied whether the expression level of the 13 differentially expressed genes, considered as candidate marker genes for T1D DKD, correlated with clinical variables contributing to DKD progression (hyperglycemia and hypertension) at or close to the time of urine collection (**Fig. 7**).

**Fig. 7:**
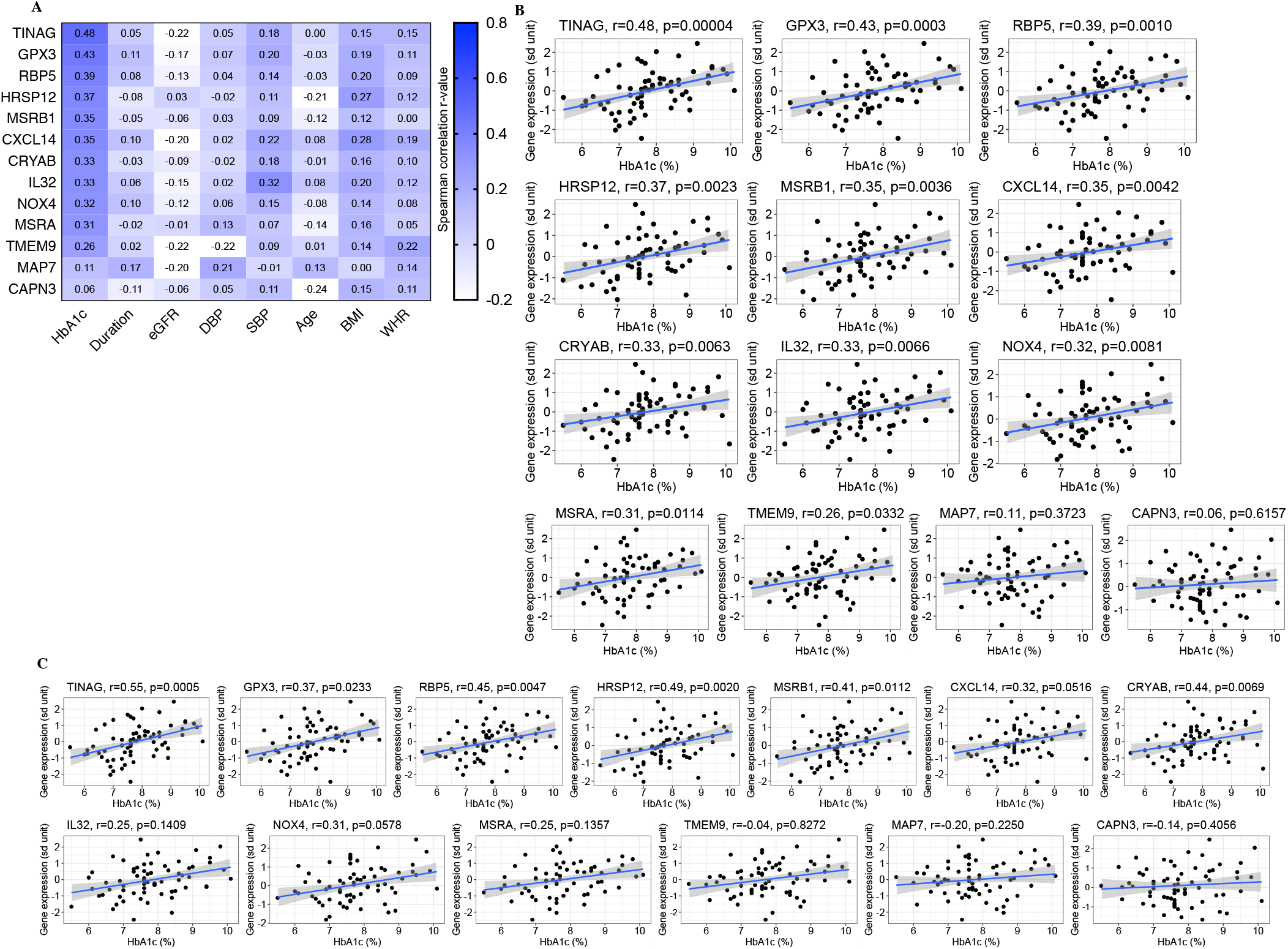
Expression level of the top differentially expressed uEV transcripts correlated with hyperglycemia. **(A)**, Heatmap depicting the strength of the correlation between expression level of the 13 differential genes in uEV with eight continuous clinical variables in the T1D individuals (n=65-67 samples, including 35-37 with normo-, 13 with micro- and 17 with macroalbuminuria). The 13 genes were obtained from differential expression analysis of uEV between macro- and normoalbuminuric T1D groups (Fig. 5). (**B** and **C)**, Correlation between expression level of the 13 genes and HbA1c level (%) in all T1D subjects (**B**, n=67), and in normoalbuminuric T1D subjects only (**C**, n=37). HbA1c was measured close to the time of urine collection. The gene expression values (log2CPM) were inverse normally transformed and converted to z score units (sd unit). All data were transformed using inverse rank transformation before Spearman’s correlation analysis (two tailed). Body mass index (kg/m^2^) (BMI); diabetes duration (Duration); diastolic blood pressure (DBP); estimated glomerular filtration rate (eGFR); glycated hemoglobin (HbA1c); log2 normalized counts per million (log2CPM); systolic blood pressure (SBP); type 1 diabetes (T1D); urinary extracellular vesicles (uEV); waist-to-hip ratio (WHR).

Most of the differentially expressed candidate genes (11 out of 13), except *MAP7* and *CAPN3*, showed a positive correlation with HbA_1c_ (**Fig. 7, A** and **B**). Of note, the strongest correlation (p≤0.001) was observed for the candidate genes known to be highly expressed in kidney (*TINAG, GPX3* and *RBP5*) compared to other human tissues (fig. S8). In a subanalysis that included samples from normoalbuminuric individuals only (from both cohorts, n=37), we also saw a trend of positive correlation between expression levels of 10 candidate genes and HbA_1c_ suggesting, that the observed correlations in the whole T1D cohort were not only driven by albuminuria or kidney disease (**Fig. 7C)**.

We next explored the origins of the differential uEV transcripts using co-expression pattern and by positioning the candidate signature on a tissue map that we created by using an external human tissue expression reference database (GTEx) (*30, 31*). Considering the urogenital organs previously established to be important contributors of uEV (**Fig. 4**), the expression level of 8 out of 12 candidate marker genes (present in GTEx dataset) of T1D DKD were higher in kidney cortex compared to prostate, bladder and testis samples in the GTEx datasets (male only, **Fig. 8A**). By pairwise correlation analysis, the co-expression pattern of the candidate genes in uEV resembled the pattern in kidney cortex when focusing on most of the kidney enriched genes (*MSRA, RBP5, GPX3, CYCL14, TINAG, NOX4, IL32* and *CRYAB*), but also in case of many of the non-kidney enriched candidate genes (*MSRB1, CAPN3, TMEM9* and *MAP7*) **(Fig. 8, B** to **E**). Further, we utilized single nuclei RNAseq data (>23,000 nuclei) from human diabetic kidney tissue (*34*), to look up the expression levels of the T1D DKD candidate genes in different cell types of the kidney. Most of the candidate genes (6 out of 9 genes that had cell type specific expression data available) showed highest expression in the proximal convoluted tubule (PCT) cells of kidney (**Fig. 8F**).

**Fig. 8:**
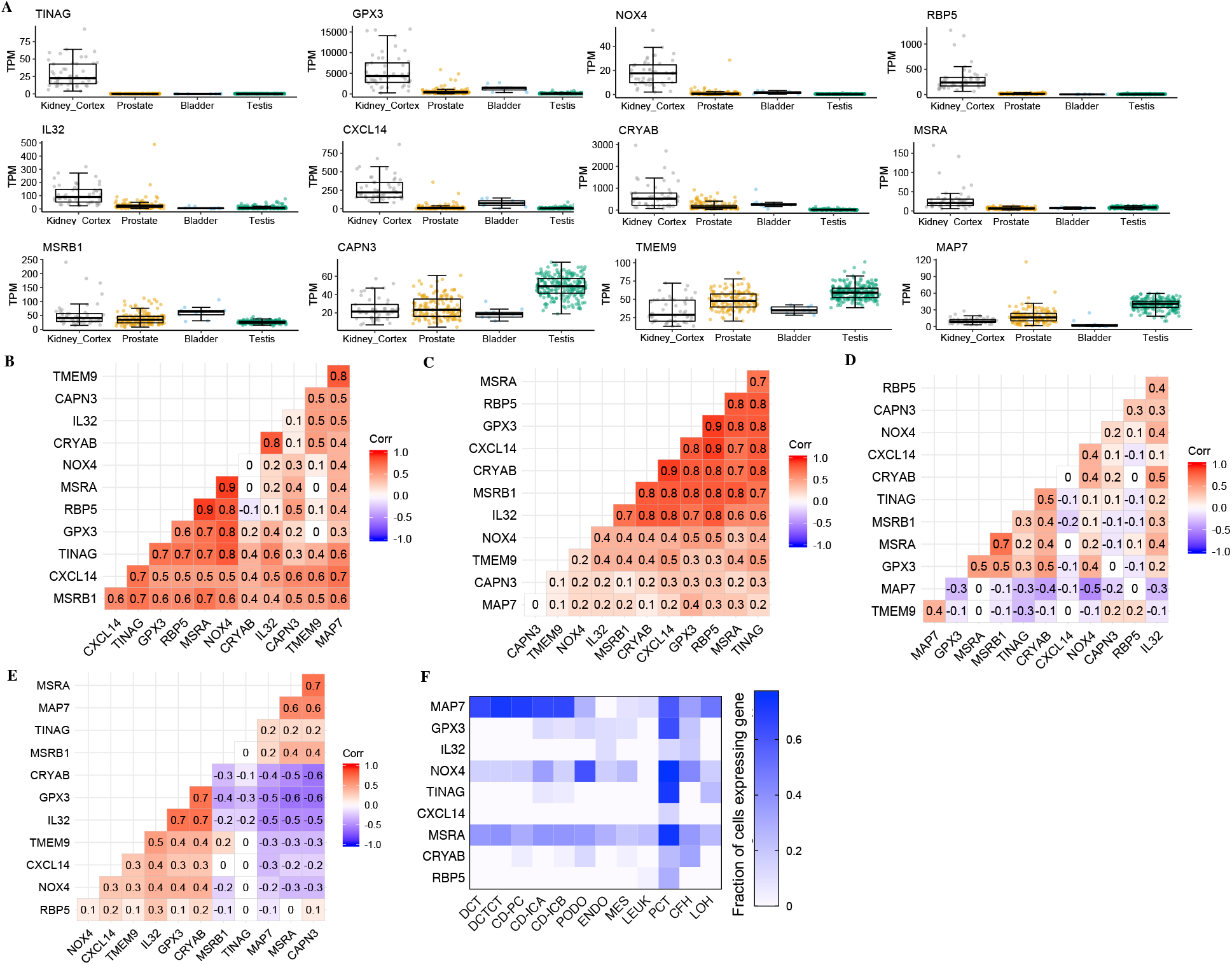
Resemblance in gene expression signature of the top DKD candidate genes between uEV and human kidney tissue. **(A)**, Expression levels (interquartile range, median and minimum/maximum summary values) of the genes-shown to be differentially expressed in uEV from macro-vs normoalbuminuric patients (12 out of 13 genes depicted)- in male urogenital organs obtained from the GTEx reference database^30,31^ (tissues from men only; kidney-cortex, n=49; prostate, n=140; bladder, n=9; testis, n=216). (**B** to **E)**, Pairwise gene expression correlation (Spearman’s correlation analysis) among the differentially expressed genes (n=12) in male urogenital organs (from GTEx, (**B)**-kidney-cortex, n=49; (**D)**-prostate, n=140; (**E)**-testis, n=216) and uEV (**C**, n=72, both T1D cohorts). (**F)**, Heatmap showing fraction of specified kidney cells expressing the nine differentially expressed genes. Data is from single cell sequencing of human diabetic kidney and obtained from Wilson PC et al. (*34*). The assessed kidney cell types were: PCT, proximal convoluted tubule; CFH, complement factor H; LOH, loop of Henle; DCT, distal convoluted tubule; CT, connecting tubule; CD, collecting duct; PC, principal cell; IC, intercalated cell; PODO, podocyte; ENDO, endothelium; MES, mesangial cell; LEUK, leukocyte. Correlation (Corr.); counts per million (CPM); diabetic kidney disease (DKD); genotype-Tissue Expression (GTEx); principal component (PC); transcripts per million (TPM); urinary extracellular vesicles (uEV).

In summary, expression analysis of candidate marker genes for T1D DKD suggested that hyperglycemia may contribute to their higher expression in uEV samples. The main contributors of the candidate marker genes to uEV could be the kidney proximal convoluted tubule cells.

### T1D DKD differential uEV mRNAs-based stress score and long-term eGFR change

All or many of the 13 differentially expressed candidate marker genes for T1D DKD showed common characteristics: 1) upregulation in DKD individuals (**Fig. 5**), 2) positive correlation with hyperglycemia (**Fig. 7**), and 3) higher expression level in kidney cortex and proximal tubular cells (**Fig. 8**). This suggested that also the underlying disease mechanism responsible for these common characteristics might be common. Based on literature regarding the 13 DKD candidate genes, we found that 6 out of 13 of the genes executed functions in cellular stress responses, namely 1) oxidative stress related responses (*GPX3, NOX4, MSRB1, MSRA*) and 2) protection of cellular proteins during stress (*HRSP12* and *CRYAB*) (table S6). In addition, the expression level of these 6 genes showed a positive correlation with HbA_1c_ (**Fig. 7**) and co-expression with each other (**Fig. 8D)**. Hyperglycemia induced stress in kidney cells including the PCT is considered to be an important disease mechanism contributing to DKD progression (*35*). Thus, we speculated that these 6 genes may be involved in hyperglycemia induced stress responses of the kidney and that their expression level in uEV could reflect the long-term change in kidney function of diabetic individuals. To test this hypothesis, we derived a stress score based on average normalized expression level of the 6 stress response related differentially expressed genes in uEV (*GPX3, NOX4, MSRB1, MSRA, HRSP12* and *CRYAB*, see methods statistics). As expected, the stress score had a clear increasing trend from normo- to macroalbuminuria and showed a positive correlation with HbA_1c_ measured at the time point of urine sample collection (**Fig. 9A** and **B**). The stress-score did not correlate with eGFR, SBP and DBP measured at the time of urine collection (**Fig. 9B)**.

**Fig. 9:**
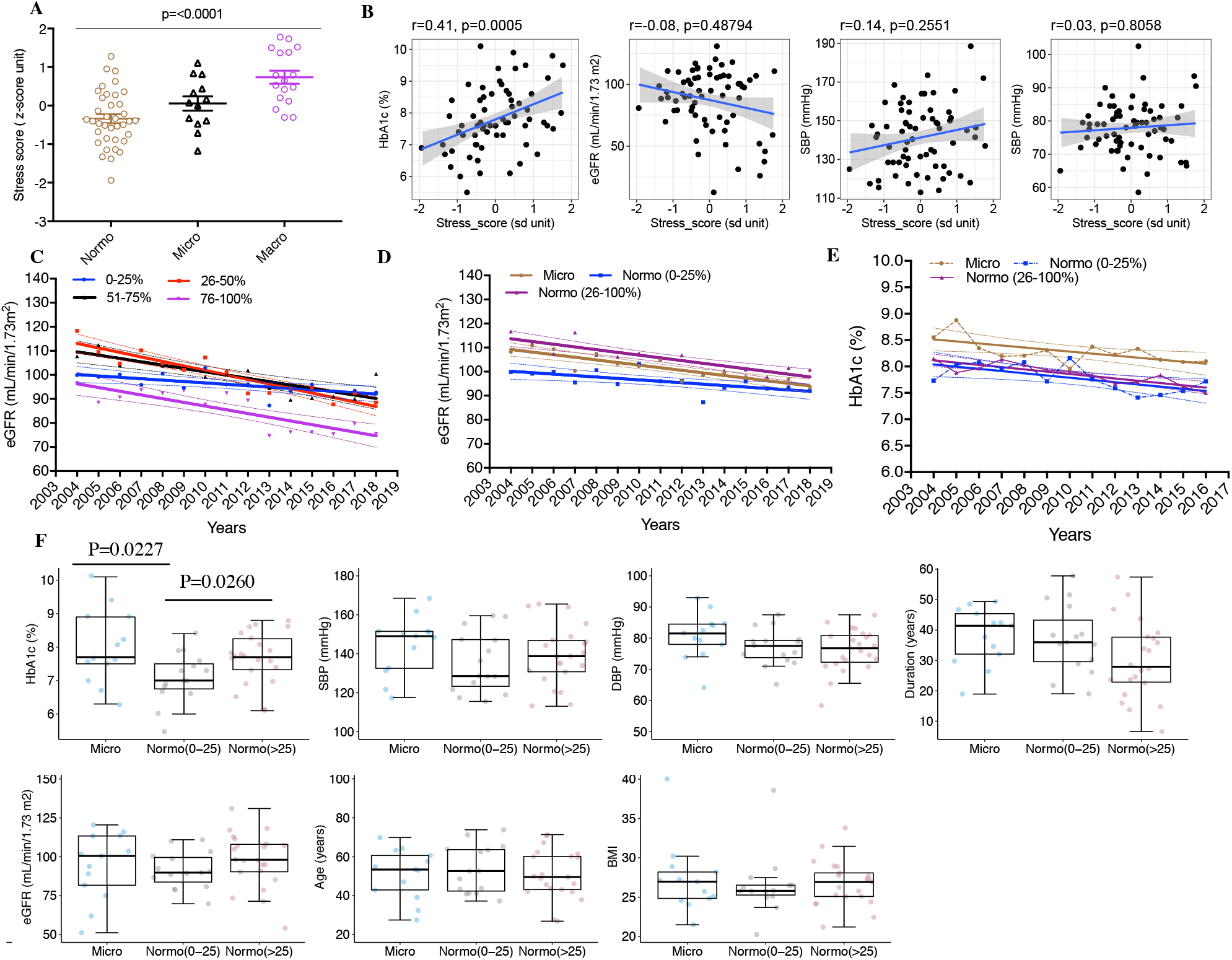
Stress score based on differential uEV mRNAs stratified T1D individuals differing in decline rate of kidney function. **(A)**, Transcriptomic stress scores in uEV from T1D individuals stratified for their albuminuria status: normo- (n=37), micro- (n=13) and macroalbuminuria (n=17). (**B)**, Correlation of HbA1c, eGFR and blood pressure (SBP and DBP) level with the transcriptomic stress score in all T1D individuals (n=67; clinical data and gene expression measured at urine collection time). **(C** and **D)**, Sequential laboratory data were retrospectively obtained from clinical databases for T1D individuals (n=51) who were stratified into quartiles based on the uEV transcriptomic stress score (n=12-13 per quartile); (**C)**, change of eGFR over 15 years (mean/year and slope) according to the transcriptomic stress score quartiles; and (**D)**, change of eGFR over 15 years in normoalbuminuric individuals within the lowest quartile (≤25%, n=11) or higher quartiles (>25%, n=19) of the stress score. For comparison, data for all microalbuminuric individuals (n=10,) with available retrospective eGFR data and without any stress score stratification are shown as a positive control; (**E)**, change of HbA1c over 13 years (mean/year and slope) as described in **D. (F)**, distribution of clinical data measured at the time of uEV collection among the microalbuminuric individuals (n=13) and normoalbuminuric individuals stratified for the stress score (lowest quartile, n=15; two middle quartiles, n=22). Data were analysed by the Kruskal-Wallis test (**A**), Spearman’s correlation analysis (two tailed, **B**), linear regression analysis (**C** to **E**) or Mann-Whitney test (**F)**. Data are in Mean ±SEM (**A)**, mean **(C** to **E**) or boxplot with interquartile range, median and summary minimum/maximum values (**F)**. Albumin excretion rate (AER); estimated glomerular filtration rate (eGFR); glycated hemoglobin (HbA1c); diabetes duration (Duration); diastolic blood pressure (DBP); macroalbuminuria (Macro); microalbuminuria (Micro); normoalbuminuria (Normo); systolic blood pressure (SBP); type 1 diabetes (T1D); urinary extracellular vesicles (uEV).

We then stratified the T1D cohorts into quartiles (0-25, 25-50, 50-75, 75-100) based on the stress score and compared annual change in clinical parameters between them over time. To compare the annual eGFR change, we collected long-term eGFR data retrospectively (15 years, from 2004 to 2018) for most of the T1D individuals (available for 51 of 67) from clinical laboratory databases. The lowest stress score quartile (0-25) showed only minimal decline in the annual eGFR (slope_(mL/min/1.73m2)_±SE; −0.58±0.19, p=0.008) (**Fig. 9C**). In contrast, the annual eGFR decline was >2-fold steeper in the highest quartile (75-100; slope_(mL/min/1.73m2)_±SE; −1.55±0.27, p=<0.0001). As the stress score was defined based on the change in macroalbuminuria group, we performed a subanalysis by excluding this group. Hence, we compared the normoalbuminuric T1D individuals within the lowest stress score quartile and within the three higher quartiles (0-25 and >25) to each other and to the group of all microalbuminuric T1D individuals (regardless of the stress score) for the decline of long-term eGFR. The normoalbuminuric group with the lowest stress score (0-25) showed only a minimal decline (slope_(mL/min/1.73m2)_±SE; −0.59±0.19, p=0.007) in the eGFR compared with normoalbuminuria group with mid-to-high stress score (>25; slope_(mL/min/1.73m2)_±SE; −1.34±0.20, p=<0.0001), which did not markedly differ from the microalbuminuria group (slope_(mL/min/1.73m2)_±SE; −1.08±0.16, p=<0.0001) (**Fig. 9D**). The overall change in the longitudinal HbA_1c_% (13 years) did not differ between the normoalbuminuria groups having low or mid-to-high stress score (0-25 and >25) (**Fig. 9E**), but the most recent HbA_1c_ level measured at the time point of uEV collection was higher for the mid-to-high (>25) stress score group similar to as observed for microalbuminuria group (**Fig. 9F**).

Overall, the analysis showed that a lower uEV stress score – a lower level of candidate gene mRNA in uEV - associates with a minimal long-term decline of kidney function. Conversely, a higher stress score associates with a faster decline of kidney function in T1D individuals regardless of their albuminuria status. This may reflect that these genes could be putative candidate genes involved in the response of the kidney to the stress caused by hyperglycemic conditions.

## Discussion

Extracellular RNAs originating from the kidney and passed to urine inside EVs could serve as a non-invasive proxy for the biopsies, a liquid kidney biopsy. However, as most EV studies have focused on small RNAs (*36*), we miss systematic characterization of whole mRNA transcriptomes in uEV, particularly with a focus on DKD. Here we characterized the uEV mRNA transcriptomes obtained from different urine collections, showed their similarity to human kidney mRNA profile and utility in discovering candidate markers for T1D DKD. This proof-of-concept study supports the idea that uEV mRNA transcriptomes capture kidney-derived gene expression signatures suitable for DKD precision medicine.

Overall, we detected expression of >10,000 genes despite focusing on the most commonly expressed genes (>90% samples) defining the most stable EV-mRNA transcriptome in population scale. The expression of large gene numbers in uEV is in line with results from some recent studies with few individuals (*23, 27, 28*). Previous studies have also confirmed the occurrence of some mRNAs in urine by qPCR even in population scale (*37*). However, why such a high number of mRNAs occur in uEV is still not known.

EVs in urine are reported to originate from the urogenital system (*38, 39*), but also from distant organs beyond the glomerular filtration barrier, including the brain (*40-42*). We detected a high proportion (>50%) of kidney enriched genes in uEV and through placing the total uEV mRNA transcriptome on a tissue map, i.e. comparing it with other human tissues, we confirmed the overall similarity with the kidney. Interestingly, irrespective of the albuminuria status, the uEV showed only a small subset of transcripts (<5%) not known to be expressed in the kidney. This suggests that kidney is the main contributor of uEV and that the glomerular filtration barrier limits the access of EV from non-urinary tract organs into the urine. Nevertheless, as we did detect some non-kidney expressed genes, we cannot totally exclude the possibility of a low degree EV transfer from circulation to urine (*43*). Out of the other urogenital system tissues of men, prostate tissue showed the closest similarity with uEV (**Fig. 4**). This finding is in line with application of uEV for screening of prostate cancer (the Exosome Diagnostic’s ExoDx Prostate cancer test) (*44*).

EV field is still in its infancy with non-standardized workflows for urine transcriptome, particularly when considering EV mRNAs as compared to miRNAs in DKD (*45*). Thus, to be able to evaluate the potential of the uEV mRNA transcriptome as a DKD biomarker source, we needed to test several pre-analytical and analytical factors. Specifically, we assessed the impact of the urine collection methods used in the clinic, 24-h or ON, which can affect the uEV concentration or cargo and thereby the detection sensitivity. We confirmed that ON urine – easier to collect than 24-h – produced almost similar mRNA profile and expression level of kidney enriched genes as the 24-h urine. Additionally, our whole uEV pipeline was reproducible, as it gave similar mRNA transcriptomes from the technical replicates processed at 1-5 months intervals through the pipeline. This suggests that urine storage at −80°C preserves uEV mRNAs well. Further, the degree of variation due to technical steps that are often assumed to cause problems, such as uEV isolation by ultracentrifugation (*46*) or sequencing in different batches (*47*), was not significant here. We also validated the mRNAseq results with qPCR for the top differential genes. We finally showed the difference in the whole uEV mRNA transcriptomes between men and women. However, the difference was minimal when focusing on kidney enriched genes. This technical information should assist in establishing a standardized workflow from biobanking of urine to future large scale uEV mRNA research.

The study revealed putative marker genes for DKD in T1D individuals. Interestingly, we found that the majority of the top differentially expressed genes between different albuminuria or eGFR status groups shared a common pattern: upregulation in T1D individuals with DKD, positive correlation with hyperglycemia, high expression in PCT and implication as cellular stress responsive genes (table S6). These top genes appear to follow the main path of the development of DKD, starting with hyperglycemia altering the metabolic flux of glucose, which causes oxidative stress mainly in PCT and further leads to inflammation, fibrosis and decline of the renal function (*48, 49*). The important roles of PCT in kidney dysfunction (DKD and CKD) is also supported by kidney tissue bulk (*9*) and single cell RNA sequencing data (*50, 34*). Remarkably, now uEV provide an opportunity to assess the expression of some PCT enriched genes (e.g. *TINAG, CXCL14 and RBP5*) highly specific for PCT, **Fig. 8**) non-invasively in T1D individuals progressing towards DKD. Based on the known functions of the candidate marker genes, it appears that many associate with kidney diseases, and our stress score genes, particularly *GPX3, HRSP12, MSRA, MSRB1* and *CRYAB*, protect the cell and/or cellular proteins during stress (table S6). They thus may have a reno-protective role by mitigating hyperglycemia-induced stress in the kidney. *GPX3* is one such antioxidant enzyme mainly produced in PCT and involved in relieving oxidative stress by neutralizing hydrogen peroxide (*51*). The *GPX3* and *MSRB1* belong to the selenoprotein class (*52*). Selenium deficiency induces oxidative stress in murine kidney cells (*53*). Recently, enhancing the *GPX3* activity by selenium supplementation was shown to rescue kidney function in T2D human subjects (*54*). *NOX4*, also one of the stress score genes, has many roles in DKD ranging from detrimental to protective and its protein level was recently shown to be decreased in PCT during CKD (*55, 56*). It is tempting to speculate that secretion of these reno-protective transcripts by PCT to urine via EVs may contribute to kidney dysfunction following hyperglycemia induced stress. Indeed, based on our clinical data, the T1D individuals with the lowest level of the stress score transcripts in uEV showed only minimal decline of the kidney function over 15 years. Thus, retaining the transcripts in the kidney may be associated with protection. Increased EV secretion during various types of stress, including oxidative stress (mechanical stretch, hypoxia, hyperglycemia, inflammation, acidosis) could contribute to the “leaking” of kidney transcripts (*57-59*). One mechanistic explanation could be that the impaired intracellular EV degradation via defective autophagy in DKD would lead to increased EV secretion (*60, 61*), as the two systems appear closely linked, which is supported by accumulating data from cell culture studies (*62-64*). Alternatively, or additionally, the observed increased transcript levels in uEV could be caused by lack of uptake and disturbed intra-renal signaling (*15, 22*). Overall, experimentation, including using EV secretion inhibitors, would help to confirm whether increased transcript secretion via uEV in T1D individuals causes a decrease in the level of reno-protective proteins in the kidney or whether it is just a consequence of an ongoing compensatory mechanism in a hyperglycemic kidney.

Among the rest of the candidate genes with no known role in cellular stress, *TINAG* associated strongly with hyperglycemia. The changes in *IL32* and *CYXCL14* suggest participation of inflammatory pathways in DKD. The expression level of *IL32* in kidney (*11*) and a genetic variant in the *TINAG* gene region have been shown to correlate with eGFR levels (rs142516820, P= 0.95×10^−13^), suggesting roles in kidney function (*65*). In summary, our hypothesis-free strategy identified DKD candidate genes, which are mostly kidney-derived and have previously established roles in kidney function.

The main limitations of our study were 1) low sample size of individual groups, 2) focusing on male subjects only (due to expected higher heterogeneity in women, which could have had an impact considering the low sample size), and 3) lack of replication of the findings in independent cohorts. However, given the novelty of the whole transcriptome EV mRNAseq methods, this is the largest study till now with inclusion of two different T1D cohorts and detailed long term retrospective clinical data. Additionally, we define a robust uEV transcriptome of >10,000 genes, show the position of uEV on tissue map, and provide evidence that uEV pipelines can be reproducible and applicable for high-impact large scale biomarker studies. Of note, better exploration of these topics has recently been warranted by both the extracellular RNA communication consortium (*36*) and Urine Task Force of the International Society for Extracellular Vesicles (*29*).

In summary, the study comprehensively shows the utility of uEV mRNA transcriptomics in capturing the gene expression patterns of the kidney and reveals type 1 DKD candidate marker genes including putative reno-protective genes. These genes could be targeted for development of novel biomarkers or therapeutic targets for DKD. Thus, we conclude that uEV offer a promising liquid kidney biopsy of DKD.

## Materials and Methods

### Study groups and ethics

Urine samples and clinical data were collected in the nation-wide prospective FinnDiane (Finnish Diabetic Nephropathy) Study targeting risk factors for DKD in type 1 diabetes (T1D), and the DIREVA study (Diabetes Registry in Vaasa), inviting all diabetic individuals living in the Vaasa Central Hospital District to participate since 2007. The studies followed the principles of the Declaration of Helsinki and all participants gave an informed consent prior to participation. The study protocol for this substudy of the FinnDiane and DIREVA studies was approved by the Ethics Committee of the Turku University Hospital. The study protocol of the FinnDiane Study has also been approved by the Ethical Committee of the Helsinki and Uusimaa Hospital District (FinnDiane 163/E5/04, 199/E5/05, 491/E5/2006; 238/13/03/00/15), and that of DIREVA, by the Ethical Committees of the Vaasa Central Hospital (6/2007) and Turku University Hospital (48/1801/2014; 116/1805/2016).

In the FinnDiane Study, type 1 diabetic men aged 18-70 years were recruited. Type 1 diabetes was defined as an onset of diabetes before the age of 40 and insulin treatment initiated within the first year after diagnosis. In spurious cases, C-peptide levels (<0.2 mmol/l) were required and medical files were reviewed. In DIREVA, male individuals with clinical type 1 diabetes aged 30-60 years with different degrees of albuminuria and no history of pancreatitis were invited. Information on diabetes heredity, history of pancreatitis or gestational diabetes was also collected to verify the diagnosis of type 1 diabetes. Study subjects were investigated at the local study centers, where trained research nurses or physicians collected information using standardized questionnaires on, among others, sex, age, age at onset of diabetes, duration of diabetes, micro- and macrovascular complication profile and current medication. The nurses measured height, weight, waist and hip circumference in light clothing, and BMI and waist-to-hip ratio (WHR) was calculated. The blood pressure was measured twice with a 5 min interval from the right arm of a sitting person after an initial rest. The mean of the two measurements was calculated and used in the analyses. Venous blood samples were taken for the assessment of HbA_1c_, blood lipids and lipoproteins (total and HDL-cholesterol and triglycerides), creatinine, fS-C-peptide, fP-Glucose, S-GAD-antibodies (DIREVA) and DNA. The samples were taken either fasting or a minimum of two hours after a light meal. In addition, retrograde data on HbA_1c_, serum and plasma creatinine for estimation of glomerular filtration rate (eGFR using the CKD-EPI formula) (*66*), urine albumin excretion rate (AER) and urine albumin-creatinine ratio (ACR) were obtained from hospital laboratory databases for the validation of the albuminuria status as well as for the longitudinal analyses.

Altogether, our study included 72 men with T1D from the two cohorts. The participants were stratified into normo-, micro- and macroalbuminuria groups. In the FinnDiane cohort, the stratification was done according to the AER (based on 24-h urine collection) in two out of three consecutive urine collections: macroalbuminuria with AER >300 mg/24-h, microalbuminuria with AER 30-300 mg/24-h, and normoalbuminuria with AER <30 mg/24-h (table S1 and S2). In the DIREVA cohort, macroalbuminuria was defined as AER (based on ON urine collection) >200 *µ*g/min or ACR (spot urine albumin/creatinine ratio) >35 mg/mmol, and microalbuminuria as AER 20-200 *µ*g/min or ACR >3.5 mg/mmol at the last clinical visit. Normoalbuminuria was defined as AER <20 *µ*g/min and ACR <3.5 (mg/mmol) at all of the clinical visits (table S1 and S2). For trajectory analysis (**Fig. 9**), we used available retrospective data on HbA_1c_ and serum or plasma creatinine from both cohorts: 1) for the last 13-15 years (available for 51 individuals), and 2) for the minimum of last five years (available for 66 individuals). For eGFR slope calculations using 15 years data, we used the average of all eGFR values per year for each individual. The classification into CKD stages (table S3) was based on retrospective data of at least > 5 years eGFR measurements for each individual (n=66), where at least two annual average values falling into the lowest eGFR range determined the CKD stage; stage 1-2 (eGFR >60), or stage 3-5 (eGFR <60). For the technical comparisons of uEV mRNAseq (**Fig. 1, 3** and fig. S5-6), we also included age and sex matched non-diabetic controls (n=4) and women with T1D and normo- or microalbuminuria (n=3) from the FinnDiane Study (table S4).

### Urine collection and isolation of uEV

The participants collected whole voids of either 24-h (FinnDiane) or ON (DIREVA) urine into containers containing Protease Inhibitor Cocktail Set III (Calbiochem, final dilution 1/1000-1/2000). The urine samples were kept in temperature ranging from ambient to +4°C during collection and delivery (the same or next day) to laboratory, where they were processed with (DIREVA) or without (FinnDiane) centrifugation at 1800g for 10 min at +4°C and frozen at −80°C. In addition, for technical comparisons, we collected similarly paired 24-h and ON urines from the same day and individuals (without centrifugation), and, 24-h urine samples from which aliquots were processed in parallel with and without centrifugation (1800g, 10 min +4°C) prior freezing (FinnDiane). EVs were isolated from 30 ml of urine by differential centrifugation as reported previously (*67, 28*). Briefly, the urine samples were vortexed for 90s, centrifuged at 8000g for 15 min at +4°C and the resulting supernatant filtered (1.2 *µ*m cellulose acetate, Whatman) to remove cell debris. EVs were pelleted by ultracentrifugation with a SW-28 rotor at 27500 rpm (136367g_max_, k-factor 254.5, Beckmann-Coulter) for 1h 30 min at +4°C, washed with PBS and the ultracentrifugation step repeated. EV pellets were suspended in PBS and stored in protein or DNA Low-bind tubes (Eppendorf, Hamburg, Germany) at −80°C.

### Western blotting, electron microscopy and nanoparticle tracking analysis of uEV

Urinary EV samples were analyzed as explained previously (*67, 28*). Briefly, Western blotting of uEV marker proteins was performed using antibodies against CD9 (SC-13118, Santa Cruz) and podocalyxin (clone 3D3, Novus Biologicals). EVs from equal volumes of urine (EVs derived from 1.7 ml of urine samples) and 20 *µ*g of protein from LNCaP cell lysate control were loaded to gels. For detection, x-ray film (Ultra Cruz™ Autoradiography Film, Santa Cruz Biotechnology (SC), Dallas, TX, USA) was used.

Electron microscopy (EM) samples were prepared with a negative staining protocol. The samples were viewed with Jeol JEM-1400 (Jeol Ltd., Tokyo, Japan) at 80 kV and imaged with Gatan Orius SC 1000B CCD-camera (Gatan Inc., USA) with 4008 × 2672 px image size and no binning.

NTA was done with Nanosight LM14 (Malvern Instruments Ltd, Malvern, UK) with blue (404 nm, 70 mW) laser and SCMOS camera (Hamamatsu photonics K.K., Hamamatsu, Japan). Samples were diluted in filtered (0.1*µ*m, Millex VV, Millipore) DPBS to obtain 40-100 particles per view. Five 30 s videos were recorded with camera level 13. The videos were analyzed using software version 3.0 (Malvern Instruments Ltd), detection threshold 5 and screen gain 10.

### RNA preparation, mRNA sequencing and data analysis

Total RNAs from the uEV were extracted using miRNEasy mini kit according to manufacturer’s instructions (Qiagen, Hilden, Germany). RNA profiles and concentrations were measured with RNA 6000 Pico Total RNA Kit run on Bioanalyzer 2100 (Agilent Technologies, Santa Clara, CA). For mRNAseq, the cDNA was prepared with SMART-Seq® v4 Ultra™ Low Input RNA Kit for Sequencing (Takara BIo Inc., Mountain View, CA) using 1 ng of total RNA as the input. The cDNA libraries were prepared with Nextera XT DNA Library Preparation and Index Kits (Ilumina Inc., San Diego, CA, USA). The cDNAs and libraries were measured with High Sensitivity D5000 ScreenTape system of TapeStation (Agilent). Paired end (150 or 300 cycles) sequencing was performed on NextSeq 550 (Illumina). RNA sequencing reads were aligned to hg38 using STAR (STAR-2.6.1d) (*68*), genome annotations were obtained from the GENCODE (Encyclopedia of Genes and Gene Variants) v227 program (*69*), and read counting was done using featureCounts (Version 1.6.4) software package (*70*) Read counts were normalized using TMM (trimmed mean of M-values) method (*71*).

### Kidney enriched gene list and GTEx analysis

We used human protein atlas to obtain the lists of 1) “kidney enriched genes” (n=413, detected in kidney and also many other tissues) with an elevated expression in the kidney compared to other tissue types, 2) “specific kidney enriched genes” (n=190) – a subset of “kidney enriched genes” (n=413) that are detected in kidney and only in some (more than one but less than one third of 37 tissues) other tissues and 3) “kidney depleted genes” (n=4631) not detected in kidney (*32, 33*). In the group of uEV samples used for technical comparisons, we detected 233 “kidney enriched genes” (≥1 count in all samples). In the two T1D cohorts combined we detected 247 “kidney enriched genes” (≥1 count in all samples) and 77 “specific kidney enriched genes”. Additionally, in both T1D cohorts we detected only 201 “kidney depleted genes” (≥1 count in all samples) in uEV. We used raw gene count (GTEx_Analysis_2017-06-05_v8_RNASeQCv1.1.9_gene_reads) and TPM values (GTEx_Analysis_2017-06-05_v8_RNASeQCv1.1.9_gene_tpm) from GTEx databases (GTEx analysis V8) for analysis (*30, 31*). For analysis purpose to focus on the main tissues, various levels of the single tissue data were merged and presented as one (e.g. sun exposed and no sun exposed skin merged and presented as “skin” only). The downstream processing of read count data used for PCA analysis were done using edgeR software (*72*).

### Taqman qPCR

Selected differentially expressed genes from mRNAseq were validated using Taqman qPCR in Fluidigm Biomark™ HD 96.96 platform (Fluidigm, San Francisco, CA, USA) according to manufacturer’s instructions. Briefly, 1,25 *µ*l of cDNAs (see preparation from RNA sequencing) were preamplified with 14 PCR-cycles according to the instructions in Fluidigm’s Real-Time PCR Analysis User Guide (PN 68000088 K1, Appendix D); the conditions were 95°C for 2 min, followed by 14 cycles of 95°C for 15 s and 60°C for 4 min. The number of preamplification cycles, 14 against 20, was tested separately with a part of the assays and samples, where 14 cycles gave better Ct values and amplification curves (data not shown). We applied one to two different assays for each gene, and 4-6 technical replicates, including two reverse transcription replicates, of each sample/assay. Water instead of sample served as a negative control. The assays with FAM-MGB labelled probes were: MAP7 (Hs01010631_m1, Hs01009609_m1), GPX3 (Hs00173566_m1, Hs01078668_m1), IL32 (Hs00170403_m1, Hs00992441_m1), NOX4 (Hs04980924_m1, Hs01379108_m1), HRSP12 (Hs01066170_m1), GAPDH (Hs99999905_m1) (all Applied Biosystems; Thermo Fisher Scientific, Inc., Waltham, MA, USA). The qPCR was performed using the online protocol “Gene Expression with the 96.96 IFC Using Fast TaqMan Assays (Biomark HD Only) (PN 100-2638 D1)” with conditions of 95°C for 60 s and 35 repeats of 97°C for 5 s and 60°C for 20 s. Data was analyzed with the method according to Livak and Schmittgen (*73*) with normalization using GAPDH. Samples with less than two successful replicas or high deltaCt (>19) were excluded from the analysis.

### Statistics

Details of statistical analysis used for each section have been described in figure legends and manuscript text. Briefly, differential expression analysis was performed based on count data using generalized linear models, as implemented in edgeR software (*72*), adjusting for age, BMI, diabetes duration and urine collection protocols (ON and 24-h) or other covariates (described in figures) as appropriate. For qPCR data analysis, deltaCt values were transformed using inverse normal transformation. Correlation analysis were performed using non-parametric Spearman’s correlation (two tailed). The transcriptional risk score was derived by averaging the inverse rank normalized expression values of the candidate genes. Slopes in the longitudinal data analysis were estimated using linear regression analysis. Data is presented as Mean ±SEM or box and whiskers plot (interquartile range, median and summary minimum/maximum values) as described in figures.

## Data Availability

All data produced in the present study are available upon reasonable request to the authors

## Acknowledgements

We acknowledge the Sequencing Units of FIMM Tech Center of the University of Helsinki, Finland, and the Department of Clinical Sciences, Diabetes and Endocrinology, Lund University, Malmö, Sweden, for RNAseq, and Biomedicum Functional Genomics Unit of the University of Helsinki, Finland, for the qPCR service, the EV Core facility of the University of Helsinki, Finland, for performing the NTA. The skillful assistance of Carola Påhls, Paula Kokko, Anna Sandelin and Jaana Tuomikangas is gratefully acknowledged. We also thank Olli Veikkola for technical assistance and Olli Kallioniemi group for laboratory facilities.

## Funding

This project has received funding from the Innovative Medicines Initiative 2 Joint Undertaking under grant agreement No 115974 (BEAt-DKD). This Joint Undertaking receives support from the European Union’s Horizon 2020 research and innovation programme and EFPIA with JDRF. The Direva Study (A.K.) has been supported by the Vasa Hospital District, State Research Funding via the Turku University Hospital, Vasa Central Hospital, Jakobstadsnejdens Heart Foundation, the Medical Foundation of Vaasa and the Finnish Medical Foundation. This substudy has also been financially supported (LG, TT) by grants from Folkhälsan Research Foundation, the Sigrid Juselius Foundation, the Academy of Finland (grants no. 263401, 267882, 312063, 336822 to LG; 312072,336826 to TT, 317599 to OPD), University of Helsinki, Ollqvist Foundation, and the Health Care Center in Vasa. The FinnDiane study was supported by grants from Folkhälsan Research Foundation, Wilhelm and Else Stockmann Foundation, Liv och Hälsa Society, Helsinki University Central Hospital Research Funds (EVO), Novo Nordisk Foundation (NNFOC0013659), and Academy of Finland (#299200 and #316664).

## Author contributions

Conceptualization: O.P.D., M.P, H.H., P-H.G, C.F, T.T., and L.G.; methodology (sample and data collection management): T.T., C.F., O.P.D., M.P., A.K., E.V. and R.B.P.; methodology (EV sample processing): M.P and K.B.; methodology (data analysis): O.P.D., M.P., K.B., C.F., and T.T.; supervision: M.P, O.P.D, C.F, T.T. and L.G.; writing (original draft): O.P.D and M.P.; writing (review and editing): T.T, A.K., C.F, K.B., O.P.D and M.P.

## Competing interests

L.G. has received research funding from Pfizer Inc, Regeneron Pharmaceuticals, Eli Lilly and Astra Zeneca. P-H.G. has received research grants from Eli Lilly and Roche; is an advisory board member for AbbVie, AstraZeneca, Boehringer Ingelheim, Cebix, Eli Lilly, Jansen, MSD, Novartis, NovoNordisk and Sanofi; and has received lecture fees from Boehringer Ingelheim, Eli Lilly, Elo Water, Genzyme, MSD, Novartis, Novo Nordisk and Sanofi. The funding sources were not involved in the design or conduct of the study.

## Data and materials availability

All data needed to evaluate the conclusions in the paper are present in the paper and/or the Supplementary Materials.

