## Supplementary tables (table S) and figures (fig. S) for "Urine extracellular vesicles capture kidney transcriptome and hyperglycemia linked mRNA signatures for type 1 diabetic kidney disease"

**This PDF file includes:**

Tables S1 to S6
Figs. S1 to S8

**Other Supplementary Materials for this manuscript include the following:**

Data S1 to Data S4

**Table S1**: Clinical characteristics of the two type 1 diabetic cohorts used in the study.

| | 24-hour collection | | Overnight collection | |
|---|---|---|---|---|
| Total Numbers (Normo/Micro/Macro) | 34 (13/7/14) | | 38 (25/8/5) | |
| | N | Mean (SEM) | N | Mean (SEM) |
| Age (years) | 30 | 50.3 (2.4) | 38 | 51.8 (2.0) |
| BMI ($kg/m^2$) | 30 | 27.9 (0.9) | 36 | 26.1 (0.5) |
| WHR | 30 | 0.96 (0.01) | 38 | 0.95 (0.01) |
| HbA1c (%) | 30 | 7.9 (0.2) | 38 | 7.7 (0.2) |
| SBP (mmHg) | 30 | 146 (3.3) | 38 | 137 (2.5) |
| DBP (mmHg) | 30 | 77 (1.6) | 38 | 79 (1.1) |
| Diabetes duration (years) | 30 | 34.0 (2.3) | 38 | 36.2 (1.6) |
| eGFR (mL/min/1.73 m2) | 30 | 81.5 (5.4) | 38 | 91.6 (3.2) |
| *AER (mg/24 hours) | 25 | 300.7 (89.7) | -- | -- |
| #AER (µg/min) | -- | -- | 37 | 38.4 (11.7) |

Normo=normoalbuminuria; Micro=microalbuminuria; Macro=macroalbuminuria; WHR=waist-to-hip ratio; HbA1c= glycated hemoglobin; SBP=systolic blood pressure; DBP= diastolic blood pressure; eGFR=estimated glomerular filtration rate; *AER= albumin excretion rate (mg/24 hours); #AER= albumin excretion rate (µg/min); N=total numbers.

**Table S2**: Clinical characteristics of the two type 1 diabetic cohorts stratified into normo-, micro- and macroalbuminuria groups that were used in global differential expression analysis.

| | Normo | | Micro | | Macro | | P value |
|---|---|---|---|---|---|---|---|
| Total Numbers | 37 | | 13 | | 17 | | |
| | N | Mean (SEM) | N | Mean (SEM) | N | Mean (SEM) | |
| Age (years) | 37 | 51 (2.2) | 13 | 51.3 (3.6) | 17 | 50.5 (2.8) | 0.9720 |
| BMI (kg/m$^2$) | 35 | 26.7 (0.6) | 13 | 27.3 (1.2) | 17 | 27.7 (1.2) | 0.8370 |
| WHR | 35 | 0.95 (0.01) | 13 | 0.93 (0.02) | 17 | 0.99 (0.02) | 0.0394 |
| HbA1c (%) | 37 | 7.4 (0.1) | 13 | 8.0 (0.30) | 17 | 8.5 (0.2) | 0.0060 |
| SBP (mmHg) | 37 | 137.6 (2.5) | 13 | 144.5 (4.1) | 17 | 147.2 (4.9) | 0.1408 |
| DBP (mmHg) | 37 | 76.6 (1.1) | 13 | 80.8 (2.0) | 17 | 78.8 (2.4) | 0.1771 |
| Diabetes duration (years) | 37 | 32.9 (2.1) | 13 | 38.1 (2.6) | 17 | 37.2 (1.9) | 0.2016 |
| eGFR (mL/min/1.73 m2) | 37 | 95.2 (2.6) | 13 | 94.0 (6.0) | 17 | 65.3 (7.6) | 0.0041 |
| *AER (mg/24 hours) | 13 | 16.3 (4.4) | 3 | 101.9 (87.7) | 9 | 777.7 (147.1) | 0.0002 |
| [#]AER (µg/min) | 24 | 5.6 (1.5) | 8 | 57.3 (12.4) | 4 | 206.7 (48.6) | <0.0001 |

P values were calculated using Kruskal-Wallis statistics.
Normo=normoalbuminuria; Micro=microalbuminuria; Macro=macroalbuminuria; WHR=waist-to-hip ratio; HbA1c= glycated hemoglobin; SBP=systolic blood pressure; DBP= diastolic blood pressure; eGFR=estimated glomerular filtration rate; *AER= albumin excretion rate (mg/24 hours); [#]AER= albumin excretion rate (µg/min); N=total numbers.

**Table S3**: Clinical characteristics of the T1D subjects stratified into groups based on chronic kidney disease (CKD) status. CKD stages were based on longitudinal eGFR data.

| | CKD stage ≤2 | | CKD stage ≥3 | | P value |
|---|---|---|---|---|---|
| Total Numbers<br>Normo/Micro/Macro | 46<br>33/9/4 | | 20<br>3/4/13 | | |
| | N | Mean (SEM) | N | Mean (SEM) | |
| Age (years) | 46 | 49.4 (2.0) | 20 | 54.0 (2.4) | 0.1698 |
| BMI (kg/m$^2$) | 46 | 26.9 (0.5) | 18 | 27.6 (1.3) | 0.9587 |
| WHR | 46 | 0.95 (0.01) | 18 | 0.98 (0.02) | 0.105 |
| HbA1c (%) | 46 | 7.6 (0.1) | 20 | 8.4 (0.3) | 0.0096 |
| SBP (mmHg) | 46 | 138.5 (2.4) | 20 | 147.1 (3.9) | 0.0768 |
| DBP (mmHg) | 46 | 78.5 (1.1) | 20 | 77.1 (1.8) | 0.5216 |
| Diabetes duration (years) | 46 | 32.1 (1.6) | 20 | 40.9 (1.7) | 0.0019 |
| eGFR (mL/min/1.73 m2) | 46 | 99.1 (2.0) | 20 | 61.1 (5.7) | <0.0001 |
| *AER (mg/24 hours) | 15 | 148.3 (103.1) | 9 | 585.4 (141.3) | 0.0035 |
| #AER (µg/min) | 28 | 20.4 (6.9) | 8 | 106.0 (42.0) | 0.0214 |

The T1D individuals (66 out of 67) were classified into chronic kidney disease stages based on eGFR data including at least > 5 years follow up. Classification required ≥2 eGFR measurements from separate hospital visits confirming the classification according to the eGFR cutoff (see Methods). The table presents the mean values of the available clinical data measured at the time of urinary EV collection. P values were calculated using Mann-Whitney test.
CKD= chronic kidney disease; Normo=normoalbuminuria; Micro=microalbuminuria; Macro=macroalbuminuria; WHR=waist-to-hip ratio; HbA1c=glycated hemoglobin; SBP=systolic blood pressure; DBP= diastolic blood pressure; eGFR=estimated glomerular filtration rate; *AER= albumin excretion rate (mg/24 hours); #AER= albumin excretion rate (µg/min); N=total numbers; T1D= type 1 diabetes.

**Table S4**: Basic clinical characteristics of the donors of the samples used in technical comparison analysis.

| Sample_ID | Sample_diabetes_status | Urine_albuminuria_status | Gender |
|---|---|---|---|
| Sam_1 | Type 1 diabetes | Microalbuminuria | Male |
| Sam_2 | Type 1 diabetes | Macroalbuminuria | Male |
| Sam_3 | Type 1 diabetes | Macroalbuminuria | Male |
| Sam_4 | Type 1 diabetes | Macroalbuminuria | Male |
| Sam_5 | Type 1 diabetes | Microalbuminuria | Male |
| Sam_6 | Type 1 diabetes | Macroalbuminuria | Male |
| Sam_7 | Type 1 diabetes | Macroalbuminuria | Male |
| Sam_8 | Type 1 diabetes | Microalbuminuria | Male |
| Sam_9 | Type 1 diabetes | Microalbuminuria | Male |
| Sam_10 | No diabetes | normoalbuminuria | Male |
| Sam_11 | No diabetes | normoalbuminuria | Male |
| Sam_12 | No diabetes | normoalbuminuria | Male |
| Sam_13 | No diabetes | normoalbuminuria | Male |

**Table S5**: Comparison of the expression levels of the top differentially expressed genes between various stratified T1D groups.

| | Normo vs Micro | | Normo vs Micro & Macro | | CKD stage ≤2 vs CKD stage ≥3 | |
|---|---|---|---|---|---|---|
| Total Numbers | 37 vs 13 | | 37 vs 30 | | 46 vs 20 | |
| | logFC | p | logFC | p | logFC | p |
| *MAP7* | 0.12 | 0.4119 | 1.14 | $9.61\times10^{-9}$ | 0.91 | $3.64\times10^{-5}$ |
| *MSRB1* | 0.28 | 0.1006 | 0.65 | $1.09\times10^{-5}$ | 0.62 | $8.67\times10^{-5}$ |
| *GPX3* | 0.49 | 0.1242 | 1.22 | $3.29\times10^{-6}$ | 1.13 | $5.30\times10^{-5}$ |
| *IL32* | 0.59 | 0.0646 | 1.28 | $3.87\times10^{-6}$ | 1.12 | $1.24\times10^{-4}$ |
| *TINAG* | 0.83 | 0.0352 | 1.34 | $7.79\times10^{-6}$ | 0.97 | $1.87\times10^{-3}$ |
| *NOX4* | 0.57 | 0.1134 | 1.45 | $1.43\times10^{-6}$ | 1.37 | $1.64\times10^{-5}$ |
| *HRSP12* | 0.23 | 0.2455 | 0.64 | $2.62\times10^{-4}$ | 0.56 | $2.35\times10^{-3}$ |
| *CAPN3* | 0.84 | 0.0961 | 1.69 | $1.58\times10^{-5}$ | 1.47 | $2.27\times10^{-4}$ |
| *CXCL14* | 0.38 | 0.2087 | 0.98 | $6.68\times10^{-5}$ | 0.81 | $1.65\times10^{-3}$ |
| *MSRA* | 0.59 | 0.0401 | 0.97 | $4.16\times10^{-5}$ | 0.72 | $3.56\times10^{-3}$ |
| *CRYAB* | 0.34 | 0.2293 | 0.90 | $1.4\times10^{-4}$ | 0.88 | $3.15\times10^{-4}$ |
| *RBP5* | 0.34 | 0.3034 | 1.02 | $1.22\times10^{-4}$ | 0.95 | $5.84\times10^{-4}$ |
| *TMEM9* | 0.49 | 0.0034 | 0.65 | $6.01\times10^{-6}$ | 0.54 | $5.57\times10^{-4}$ |

The two T1D cohorts were stratified based on albuminuria and chronic kidney disease stages based on eGFR data as shown in Supplementary Table 2 and 3 respectively. In the table, we present comparisons of the top 13 differentially expressed genes (obtained from normo vs macro comparison, as shown in Fig. 5), between the T1D groups as indicated.
The differential gene expression analysis was performed based on count data using generalized linear models adjusting for age, body mass index, diabetes duration and urine collection protocols (overnight and 24 hour). Fold change is presented in log10 FC scale.
CKD= chronic kidney disease; FC, fold change; eGFR=estimated glomerular filtration rate; Normo=normoalbuminuria; Micro=microalbuminuria; Macro=macroalbuminuria; T1D= type 1 diabetes.

**Table S6**: Stress response -related functions and association to kidney diseases for the proteins coded by the differentially expressed uEV transcripts used in the construction of the stress score.

| Stress score gene | Name | Biological funtion of proteins related to stress response , Reference | Association with the kidney diseases strength: -log10 (p-value) |
|---|---|---|---|
| *CRYAB* | Crystallin Alpha B | Chaperone function<br>Response to hydrogen peroxide and hypoxia<br>https://www.uniprot.org/uniprot/P02511 | 2.5 |
| *GPX3* | Glutathione Peroxidase-3 | Glutathione peroxidase activity<br>Cellular response to oxidative stress<br>Hydrogen peroxide catabolic process<br>Response to lipid hydroperoxide<br>Selenoprotein<br>https://www.uniprot.org/uniprot/P22352 | 2.5 |
| *NOX4* | NAD(P)H oxidase 4 | Oxygen sensor activity<br>Superoxide-generating NAD(P)H oxidase activity<br>Cellular response to oxidative stress<br>Reactive oxygen species metabolic process<br>Response to hypoxia<br>https://www.uniprot.org/uniprot/Q9NPH5 | 2.5 |
| *MSRB1* | Methionine Sulfoxide Reductase B1 | Peptide-methionine (R)-S-oxide reductase activity during Oxidative stress.<br>Protein repair<br>Selenoprotein<br>https://www.uniprot.org/uniprot/Q9NZV6 | 1.8 |
| *MSRA* | Methionine Sulfoxide Reductase | Oxidoreductase<br>Protein repair<br>Cellular response to oxidative stress<br>https://www.uniprot.org/uniprot/Q9UJ68 | 1.8 |
| *HRSP12* | Heat-responsive protein 12 | Hydrolase<br>2-iminobutanoate deaminase activity<br>May facilitate the release of ammonia from potentially toxic reactive metabolites, reducing their impact on cellular components.<br>https://www.uniprot.org/uniprot/P52758 | 1.6 |

The information of molecular function and biological pathway related to stress responses were extracted from UniProt protein data base (http://www.uniprot.org). The information regarding association with kidney diseases were obtained from the Harmonizome database (https://maayanlab.cloud/Harmonizome/gene_set/Kidney+Diseases/CTD+Gene-Disease+Associations) (https://doi.org/10.1093/database/baw100).
In the table, the stress score candidate genes were ordered based on their association strength with the kidney diseases in the Harmonizome database. uEV, urinary extracellular vesicles.

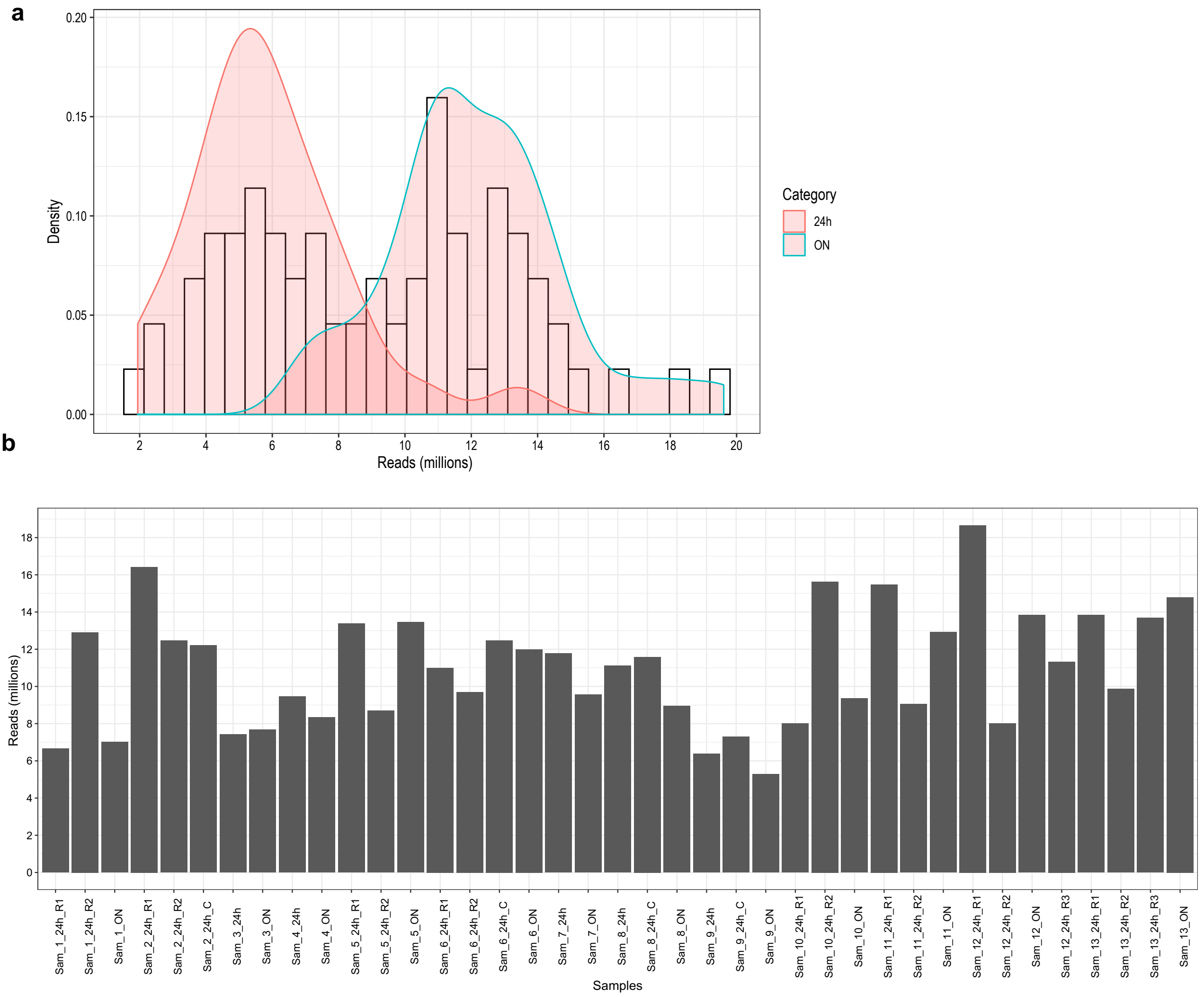


**Fig. S1: Number of raw unmapped sequencing reads from uEV mRNA sequencing in the whole T1D study cohort and technical samples.**

**(a),** Distribution of read numbers (density plot) in the whole study cohort consisting of 24h (n=34) and overnight (n=38) urine collections from T1D samples. (**b**), Number of raw sequencing reads obtained from various technical study samples (n=13 unique samples) used in technical comparisons.

Data (read numbers/density) for 9 samples overlap between **a** and **b.** These 9 samples in technical comparison analysis (Sam_1_24h_R1, Sam_2_24h_R1, Sam_3_24h, Sam_4_24h, Sam_5_24h_R1, Sam_6_24h_R2, Sam_7_24h, Sam_8_24h, Sam_9_24h) were from T1D cohorts (as depicted in **a**) that have been used as reference in their other respective pair comparison analysis.

C, centrifuged; 24h, 24 hour; ON, overnight; R, replicate; T1D, Type 1 diabetes, uEV, urinary extracellular vesicles.

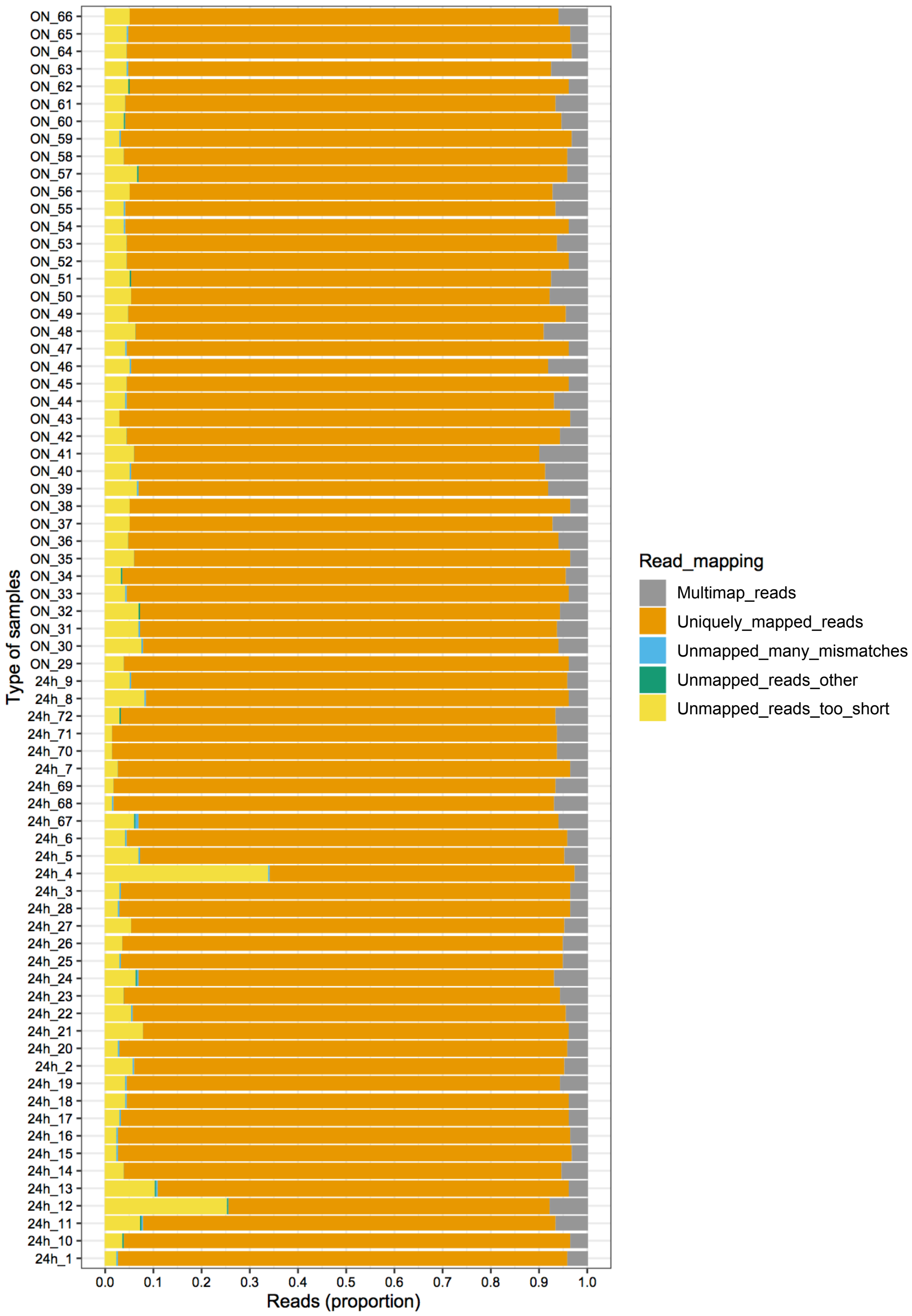


**Fig. S2: Mapping  of raw sequecing reads from uEV mRNA sequencing to human genome in the whole T1D study cohort.**

Figure shows the proportions of uniquely-mapped, multi-mapped and unmapped uEV mRNA sequencing reads to human genome in each sample. Alignments of the raw sequecing reads to human genome were done using STAR (68).
24h, 24 hour; ON, overnight; T1D, Type 1 diabetes, uEV, urinary extracellular vesicles.

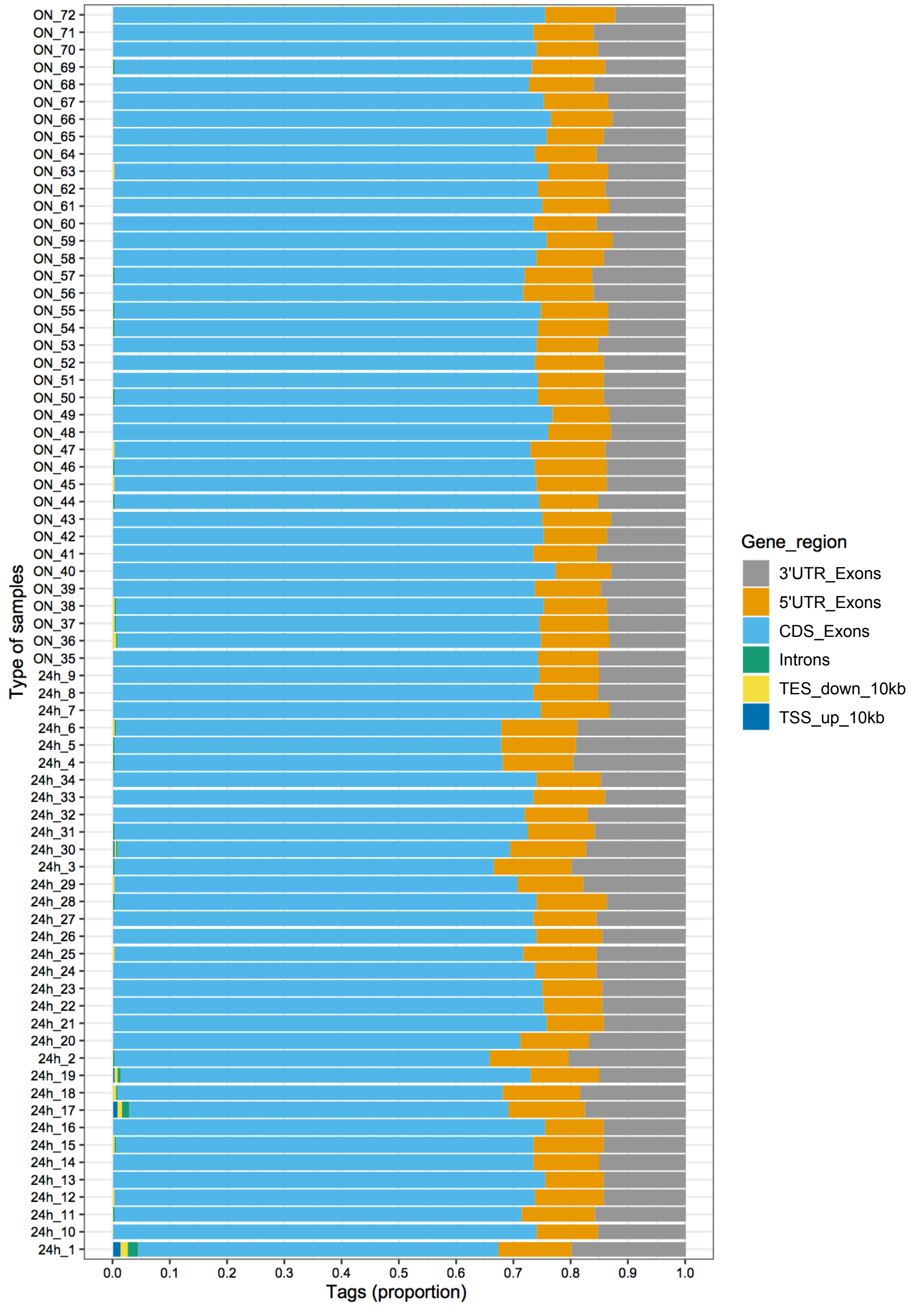


**Fig. S3: Mapping distribution of uniquely mapped mRNA sequecing reads from uEV around the coding regions of human genome in the whole T1D study cohort.**

Figure shows the proportions of tags mapping to exons, 3' or 5' UTR, introns or intergenic parts i.e. TSS or TES 10 kb (10 kb upstream of TSS or downstream of TES) in each sample. Alignments of the raw sequecing reads to human genome were done using using STAR (68).
CDS, coding sequence; ON, overnight; TSS, transcriptional start site; TES, transcriptional end site; T1D, Type 1 diabetes, uEV, urinary extracellular vesicles; UTR, untranslated region; 24h, 24 hour; ON, overnight.

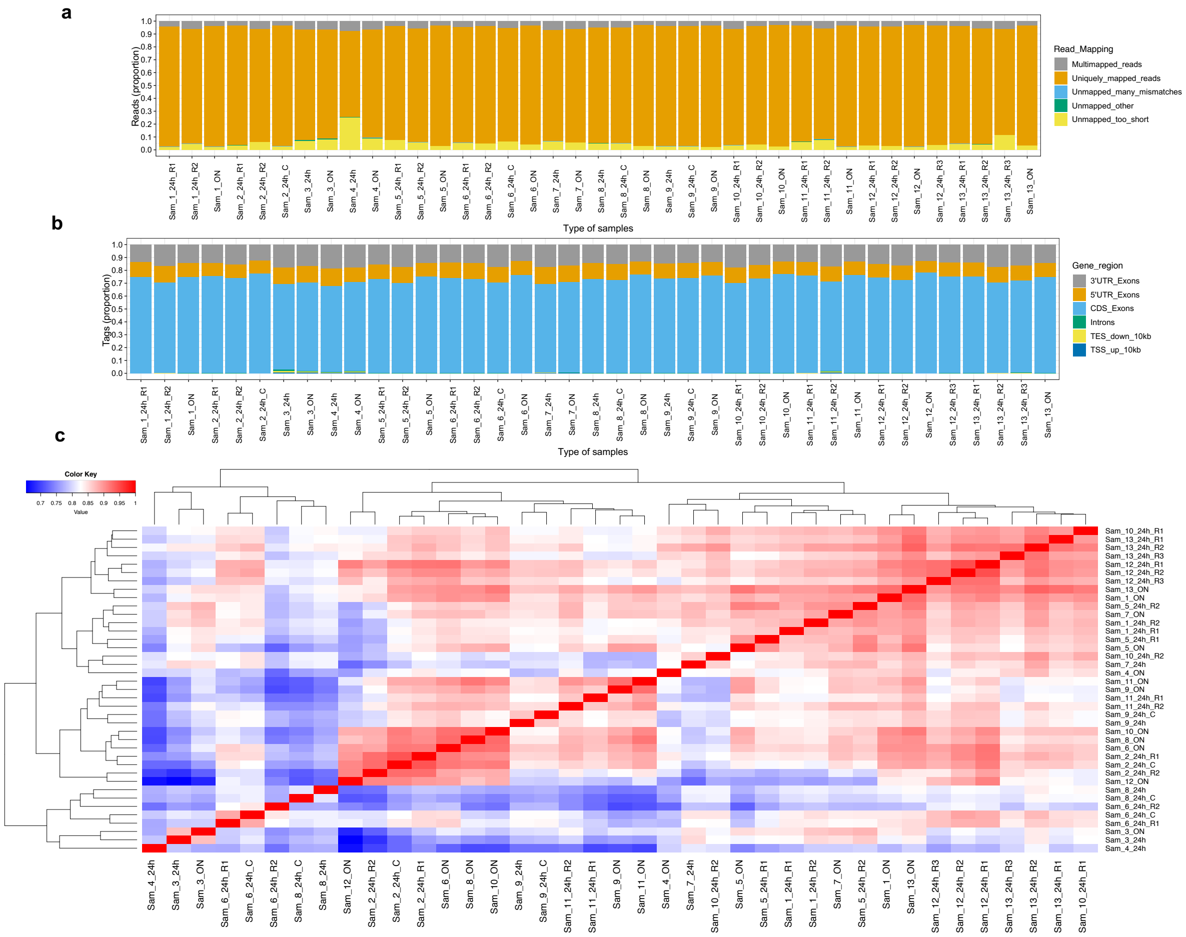


**Fig. S4 . Mapping distributions and correlation of the mRNA sequencing reads in the uEV samples used for technical comparisons.**

**(a)**, Proportions of uniquely mapped, multi-mapped and unmapped raw sequencing reads.

**(b)**, Proportions of tags mapping to exons, 3’ or 5’ UTR, and introns or intergenic parts i.e. TSS or TES 10 kb (10 kb upstream of TSS or downstream of TES).

**(c)**, Sample-to-sample correlation heatmaps and hierarchical clustering based on count data (n= 10,449 genes). Technical sample comparisons included: 1) Technical replicates; aliquots of individual samples (24 hour urine collection) were processed through the uEV mRNA sequencing pipeline twice (n=6 donors, T1D: Samples 1, 2, 5 and 6; Non-diabetic: Samples 10 and 11) or thrice (n=2 non-diabetic donors: Samples 12 and 13) at 1-5 months intervals. 2) Centrifugation test samples; aliquots of individual urine samples (24 hour collection) were processed with and without centrifugation before freezing (n=4 T1D donors: Samples 2, 6, 8 and 9). 3) Samples from overnight vs 24 hour urine collections; samples from individual donors were collected from the same day (n=12 donors, T1D: Samples 1, 3-9; Non-diabetic: Samples 10-13).

Data for 9 samples in **a**, **b** overlap with Fig. 2 and 3 respectively. These 9 samples in technical comparison analysis (Sam_1_24h_R1, Sam_2_24h_R1, Sam_3_24h, Sam_4_24h, Sam_5_24h_R1, Sam_6_24h_R2, Sam_7_24h, Sam_8_24h, Sam_9_24h) were from T1D cohorts (included in Fig. 2 and 3) that have been used as reference in their other respective pair comparison analysis. Alignments of the raw sequecing reads to human genome were done using using STAR (*68*). Correlation analysis was performed using non-parametric Spearman’s (two-tailed) method.

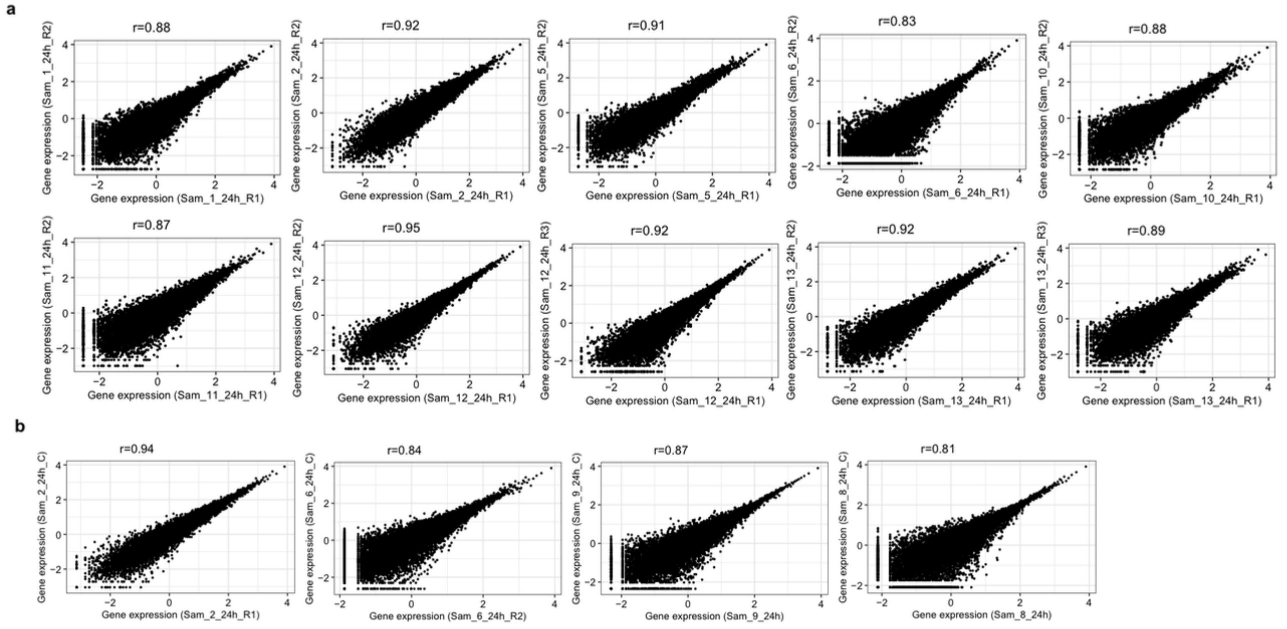


**Fig. S5: Gene expression distributions between the technical replicate pairs of uEV samples.**

Distribution of expression levels of all common protein coding genes (n=10,449 genes detected in > 90% of all technical samples) in (**a**), technical replicates - aliquots of individual samples (24 hour urine collection) were processed through the uEV mRNA sequencing pipeline twice (n=6 donors, T1D: Samples 1, 2, 5 and 6; Non-diabetic: Samples 10 and 11) or thrice (n=2 non-diabetic donors: Samples 12 and 13) at 1-5 months intervals and (**b**), Centrifugation test samples- aliquots of individual urine samples (24 hour collection) were processed with and without centrifugation before freezing (n=4 T1D donors: Samples 2, 6, 8 and 9).
All samples were from 24-hour (24h) urine collections. C, centrifuged; Sam, sample; R, Replicate; T1D, Type 1 diabetes; uEV, urinary extracellular vesicles.
Original gene expression levels (log2CPM) were inverse normally transformed and converted to z score unit (sd unit). Correlation analysis (r) was performed using non-parametric Spearman's (two-tailed) method, P<0.0001.

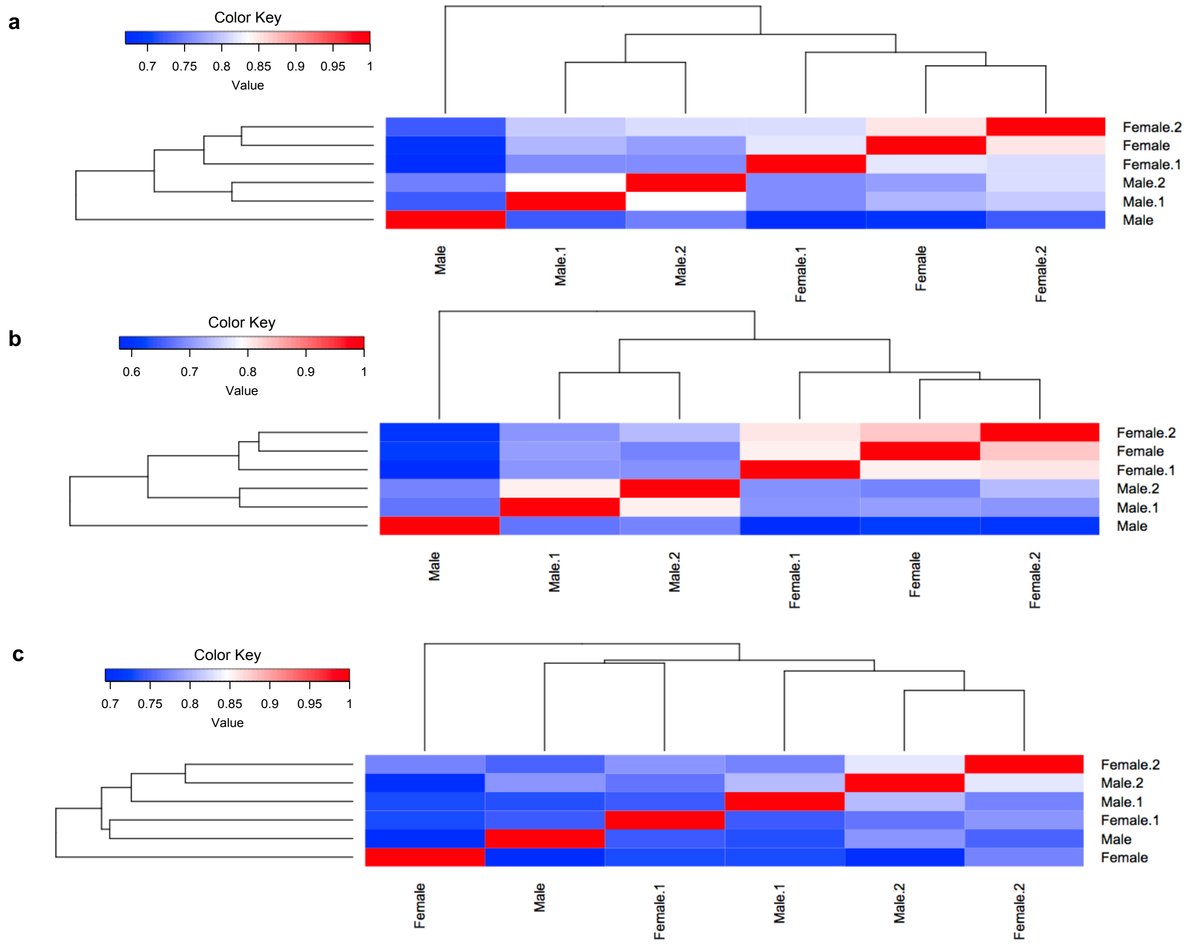


**Fig. S6: Comparison of uEV mRNA profiles between men and women.**

The heatmaps show sample-to-sample correlation (Spearman's) and hierarchical clustering based on the expression levels of (**a**)- whole uEV mRNA transcriptome (n=10, 596 genes) or (**b**)- a subset of sex chromosome genes (chromosome X and Y, n=329 genes) or (**c**)- a subset of kidney-enriched genes (n=247, based on Human protein Atlas, see Methods). Analysis included three samples from T1D female donors and three samples from T1D male donors matching for the albuminuria status of the women. All samples were from 24-hour urine collections. T1D, Type 1 diabetes; uEV, urinary extracellular vesicles.

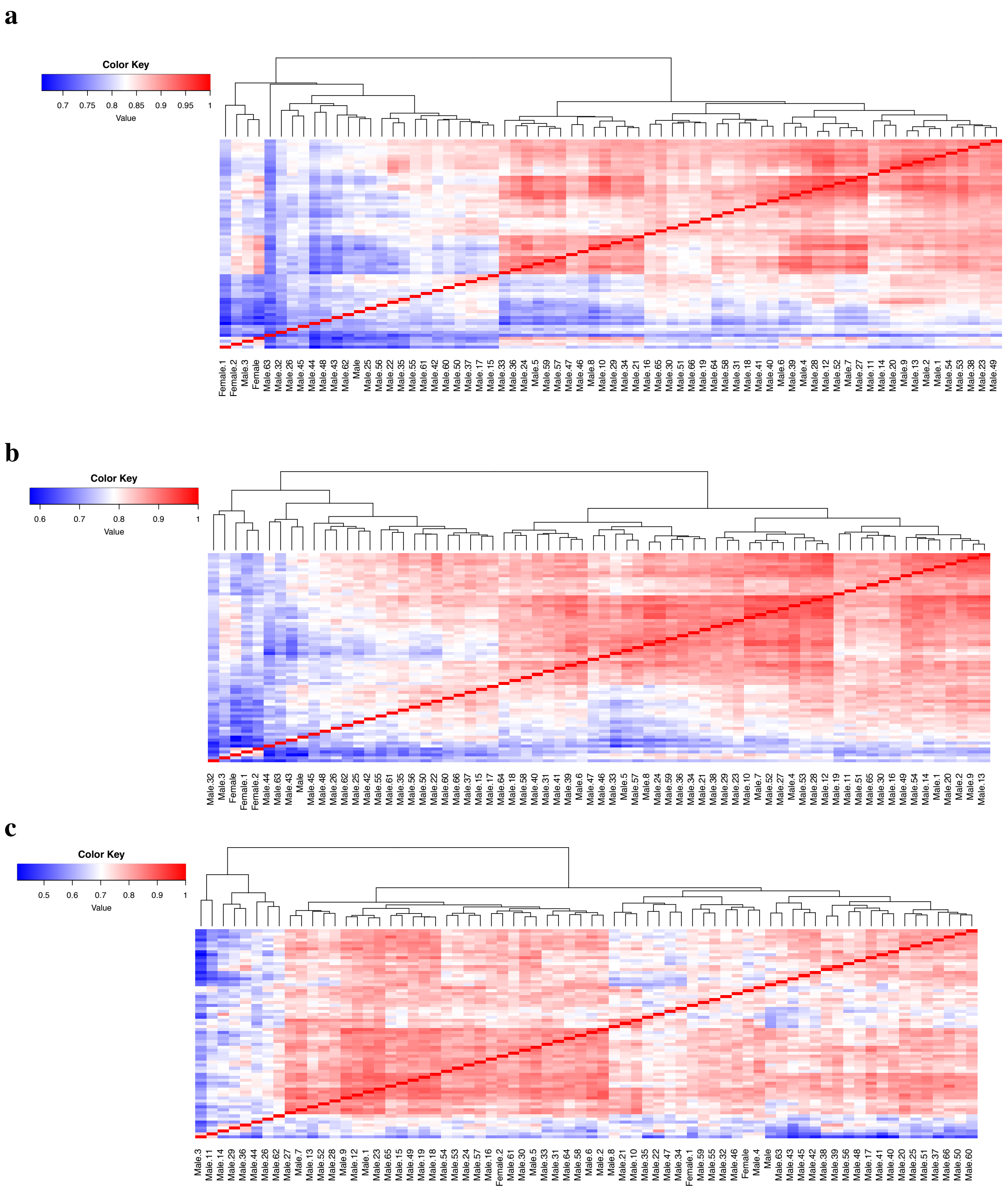


**Fig. S7: Comparison of uEV mRNA profiles between men and women.**

The heatmaps show sample-to-sample correlation (Spearman's) and hierarchical clustering based on the expression levels of (**a**)- whole uEV mRNA transcriptome (n=10, 596 genes) or (**b**)- a subset of sex chromosome genes (chromosome X and Y, n=329 genes) or (**c**- a subset of kidney-enriched genes (n=247, based on Human protein Atlas, see Methods). Analysis included three samples from T1D female donors (24-hour urine collection) and 67 samples from T1D male donors (24-hour and overnight urine collections). T1D, Type 1 diabetes; uEV, urinary extracellular vesicles.

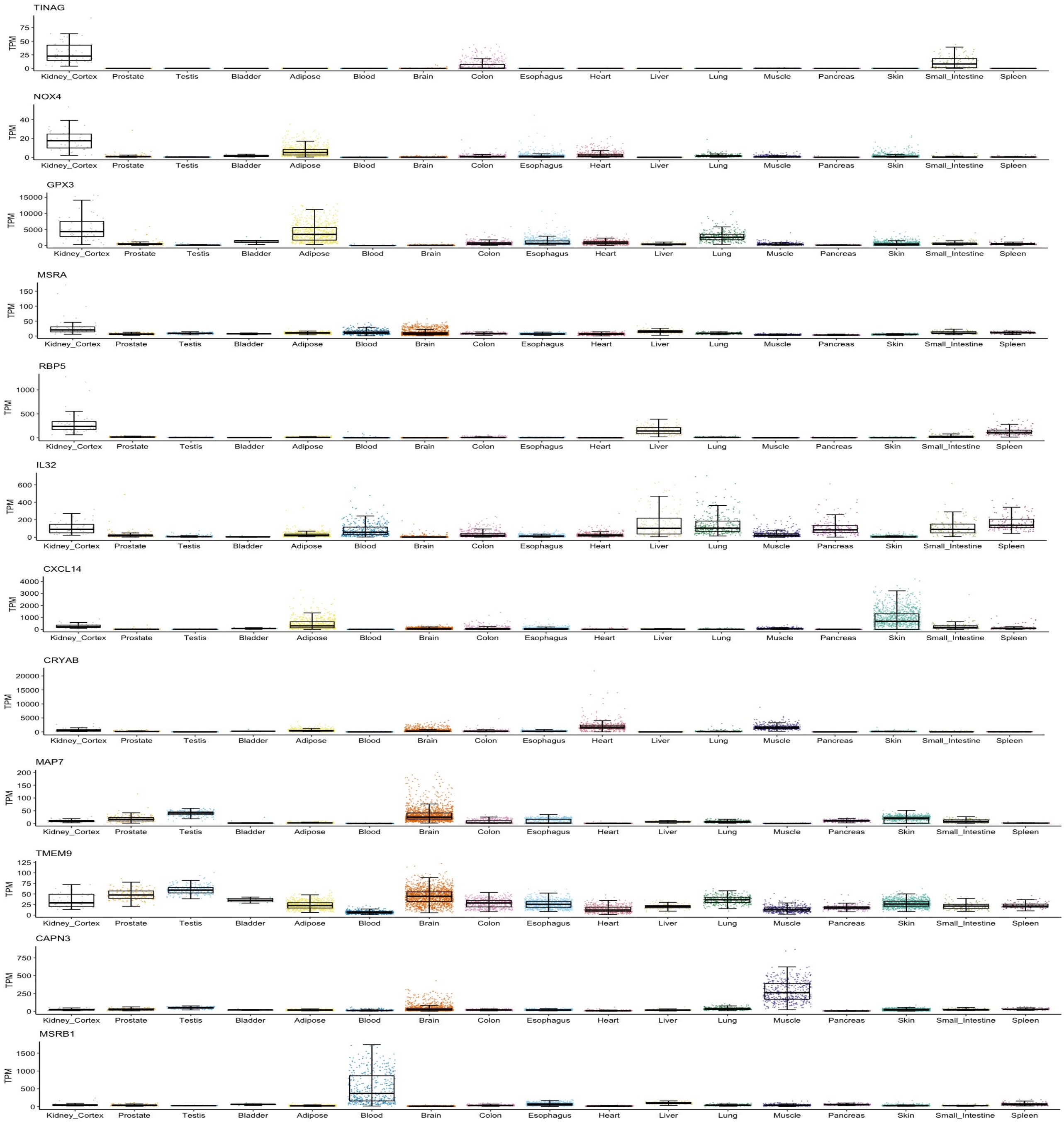


**Fig. S8: Gene expression levels of the top differentially expressed genes in various human tissues.**

Figure depicts the gene expression levels of 12 differentially expressed candidate markers for type 1 DKD in various human tissues. Differentially expressed genes were derived from comparison of uEV between normoalbuminuria and macroalbuminuria groups. Data was extracted from the GTEx reference database (*30*, *31*), using tissues derived from men only (n, Adipose=692, Bladder=9, Blood=426, Brain=1612, Colon=475, Esophagus=866, Heart=515, Kidney Cortex=49, Liver=138, Lung=327, Muscle=474, Pancreas=189, Prostate=140, Skin=1088, Small Intestine=118, Spleen=132, Testis=216). The boxplots depict the expression levels in TPM scale as extracted from

GTEx database including the interquartile range, median and summary minimum/maximum values. DKD, diabetic kidney disease; TPM, transcripts per million; uEV, urinary extracellular vesicles.
